# CRISIS AFAR: An International Collaborative Study of the Impact of the COVID-19 Pandemic on Youth with Autism and Neurodevelopmental Conditions

**DOI:** 10.1101/2022.04.27.22274269

**Authors:** Bethany Vibert, Patricia Segura, Louise Gallagher, Stelios Georgiades, Panagiota Pervanidou, Audrey Thurm, Lindsay Alexander, Evdokia Anagnostou, Yuta Aoki, Catherine S.N. Birken, Somer L. Bishop, Jessica Boi, Carmela Bravaccio, Helena Brentani, Paola Canevini, Alessandra Carta, Alice Charach, Antonella Costantino, Katherine T. Cost, Elaine Andrade Cravo, Jennifer Crosbie, Chiara Davico, Alessandra Gabellone, Federica Donno, Junya Fujino, Cristiane Tezzari Geyer, Tomoya Hirota, Stephen Kanne, Makiko Kawashima, Elizabeth Kelley, Hosanna Kim, Young Shin Kim, So Hyun (Sophy) Kim, Daphne J. Korczak, Meng-Chuan Lai, Lucia Margari, Gabriele Masi, Lucia Marzulli, Luigi Mazzone, Jane McGrath, Suneeta Monga, Paola Morosini, Shinichiro Nakajima, Antonio Narzisi, Rob Nicolson, Aki Nikolaidis, Yoshihiro Noda, Kerri Nowell, Miriam Polizzi, Joana Portolese, Maria Pia Riccio, Manabu Saito, Anish K. Simhal, Martina Siracusano, Stefano Sotgiu, Jacob Stroud, Fernando Sumiya, Ida Schwartz, Yoshiyuki Tachibana, Nicole Takahashi, Riina Takahashi, Hiroki Tamon, Raffaella Tancredi, Benedetto Vitiello, Alessandro Zuddas, Bennett Leventhal, Kathleen Merikangas, Michael P Milham, Adriana Di Martino

**Author notes:** **Corresponding Author:** Adriana Di Martino, MD, Autism Center, Child Mind Institute, 101 E 56th Street, Third Floor, New York, NY 10022, USA,. equal contribution (Bethany Vibert, Patricia Segura).

## Abstract

**Importance:** Heterogeneous mental health outcomes during the COVID-19 pandemic are recognized in the general population, but it has not been systematically assessed in youth with neurodevelopmental disorders (NDD), including autism spectrum (ASD).

**Objective:** Identify subgroups of youth with ASD/NDD based on the pandemic impact on symptoms and service changes, as well as predictors of outcomes.

**Design, Setting, and Participants:** This is a naturalistic observational study conducted across 14 North American and European clinical and/or research sites. Parent responses on the Coronavirus Health and Impact Survey Initiative (CRISIS) adapted for Autism and Related Neurodevelopmental Conditions (AFAR) were cross-sectionally collected from April to October 2020. The sample included 1275, 5-21 year-old youth with ASD and/or NDD who were clinically well-characterized prior to the pandemic.

**Main Outcomes and Measures:** To identify impact subgroups, hierarchical clustering analyzed eleven AFAR factors measuring pre- to pandemic changes in clinically relevant symptoms and service access. Random forest classification assessed the relative contribution in predicting subgroup membership of 20 features including socio-demographics, pre-pandemic service, and clinical severity along with indices of COVID-19 related experiences and environments empirically-derived from AFAR parent responses and global open sources.

**Results:** Clustering analyses revealed four ASD/NDD impact subgroups. One subgroup - *broad symptom worsening only* (20% of the aggregate sample) - included youth with worsening symptoms that were above and beyond that of their ASD/NDD peers and with similar service disruptions as those in the aggregate average. The three other subgroups showed symptom changes similar to the aggregate average but differed in service access: *primarily modified services* (23%), *primarily lost services* (6%), and *average services/symptom changes* (53%). Pre-pandemic factors (e.g., number of services), pandemic environments and experiences (e.g., COVID-19 cases, related restrictions, COVID-19 Worries), and age emerged in unique combinations as distinct protective or risk factors for each subgroup. Together they highlighted the role of universal risk factors, such as risk perception, and the protective role of services before and during the pandemic, in middle childhood.

**Conclusions and Relevance:** Concomitant assessment of changes in both symptoms and services access is critical to understand heterogeneous impact of the pandemic on ASD/NDD youth. It enabled the delineation of pathways to risk and resilience that include universal and ASD/NDD specific contributors.

## INTRODUCTION

Pediatric populations are vulnerable to the sudden and pervasive disruptions in their daily life, such as those brought by the COVID-19 pandemic.^1,2^ Those with neurodevelopmental disorders (NDD) have been identified by parents, educators, clinicians, and policy makers, as requiring specific attention due to the range of preexisting behavioral, emotional, and learning difficulties.^3–5^ Empirical reports support this notion highlighting disruptions in ongoing care^6–9^ and increases in behavioral and emotional difficulties in youth with NDD since the start of the pandemic.^9–21^ Here, we report our international effort to assess heterogeneity in the impact of the COVID-19 pandemic and predictors of outcomes in previously well-characterized youth with autism spectrum disorder (ASD) and/or other NDD.

The focus on variable outcomes is motivated by prior clinical and disaster research. Clinical literature indicates that ASD/NDD are highly heterogeneous in symptom presentations, comorbidities, intellectual abilities, and adaptive functioning.^22–24^ Disaster research, before and during the COVID-19 pandemic, has shown that the degree of severity of prior mental illness, disaster exposure, and perceived risk, are predictors of negative outcomes.^19,21,25–27^ A comprehensive understanding of the contribution of both disaster- and clinically-related predictors of outcomes in youth with ASD/NDD is needed to inform recovery efforts and prepare for future crises.

Towards this goal, we adapted the Coronavirus Health and Impact Survey Initiative (CRISIS)^27^ for Autism and Related Neurodevelopmental Conditions (AFAR). CRISIS was designed to capture the multifaceted nature of risk during the COVID-19 pandemic in the general population by quantitatively assessing life changes, perceived risk, and worries about COVID-19, as well as mental health before and during the pandemic. Previous work in the general population established CRISIS psychometrics and feasibility in delineating distinct life stress profiles and their predictive role in mental health outcomes.^27,28^ While preserving the original structure of CRISIS, AFAR aimed to quantify changes in domains affected by or known to impact changes in daily life and therapeutic services in ASD/NDD. To investigate the contributions of pandemic-related aspects, such as COVID-19 case rates and restrictions, and optimize the balance between sample size and characterization, we formed a collaborative international network of investigators aiming to collect AFAR surveys from previously well-characterized youth with ASD/NDD in a naturalistic observational framework. Using multivariate data-driven analyses in a cross-sectional AFAR dataset of N=1275 individuals, we identified distinct subgroups of impact and their predictors.

## METHODS

### Survey Development

A workgroup of ASD/NDD experts (ADM, LG, SG, PP, AT, BV) led the adaptation from the CRISIS Parent/Caregiver Baseline Form. The adaptation included assessments of clinical domains relevant to ASD/NDD and services, while maintaining the existing structure of CRISIS.^27,29^ Empirical evidence on the impact of disasters on ASD/NDD was limited to one study reporting worsening in adaptive funcitoning.^30^ Therefore, along with adaptive skills, we prioritized the assessment of symptoms known to be affected by or to impact adjustment. These encompassed restricted and repetitive behaviors/interests (RRB)^31–33^ externalizing and internalizing symptoms that often co-occur in ASD/NDD.^22,24^ Parent/caregiver questions were developed to target observable behaviors rather than reporting internal states. To contain the survey length, the CRISIS Mood State and Substance Use domains were dropped. Like CRISIS, symptoms were rated on a Likert-scale based on three months prior to the COVID-19 start in the respondent’s geographical area and over the two weeks prior to completion (*Prior* and *Current* time points, respectively).

To evaluate changes in service access, we derived items from a survey developed during the pandemic for people with syndromic intellectual disabilities and their caregivers.^8^ Questions queried changes in therapeutic services typically received both within and outside school settings in the respondents geographical area, following the start of the pandemic. The remainder of the original CRISIS was unchanged, except for some rewordings or additional response options (e.g., sleep problems), as detailed in eFigure1 and Methods in eAppendix.

Like CRISIS, questions were developed for individuals aged five to 21 years; a later review identified a subset of questions developmentally applicable for ages as young as three years (eFigure 1, Supplementary Methods). The initial adaptation was developed in English and then translated into five other languages and updated with rewordings in consultation with the larger AFAR network. The final version 0.5.1 of AFAR Parent/Caregiver Baseline Form (3-21) included 96 independent items, with 34 questions asked twice for *Prior* and *Current* timepoints; AFAR is freely available for use by other investigators.^29^

### Data Collection

AFAR Parent/Caregiver Baseline surveys were collected in 15 samples at 14 research and/or clinical institutions in Europe and North America. Data were collected cross-sectionally over the first six months of the pandemic (April-October 2020; Figure1A, eTable1, Methods in eAppendix). Parents of children aged three to 21 years with previously established clinician-based DSM-IV/5^34,35^ or ICD-10^36^ diagnosis of ASD and/or other NDD were invited to complete AFAR. Along with diagnostics, when feasible, previously collected information on intellectual functioning and symptoms were shared. IRB approval for collection and sharing of de-identified AFAR and related data was obtained at each institution.

**Figure 1.**
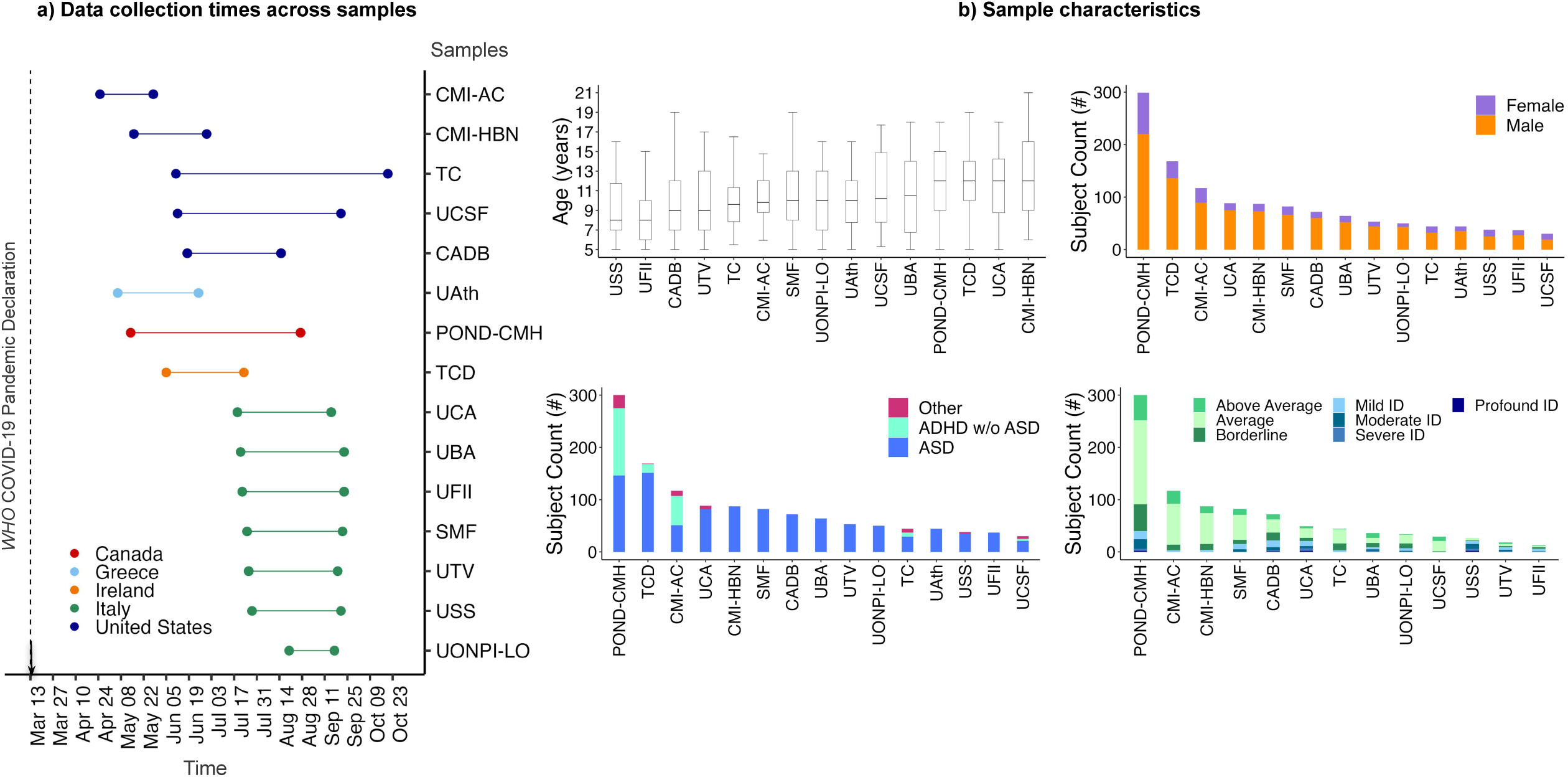
Data Collection Time Periods and Characteristics. a) Data Collection Times for each contributing samples (a) and their characteristics (b) Abbreviations: CMI-AC=Child Mind Institute-Autism Center; CMI-HBN=CMI-Healthy Brain Network; TC=Thompson Center; UCSF=University of California San Francisco; CADB=Center for Autism and Developing Brain, Weill Cornell Medical College/New York Presbyterian Hospital; UAth=University of Athens, National & Kapodistrian University of Athens, School of Medicine, First Department of Pediatrics, Unit of Developmental and Behavioral Pediatrics. “Aghia Sophia” Children’s Hospital; POND-CMH=Province of Ontario Neurodevelopmental Network, COvid Mental Health collaboration; TCD=Trinity College Dublin; UCA=University of Cagliari, Child & Adolescent Neuropsychiatry Unit, A.Cao Paediatric Hospital; UBA=University Bari, Child Neuropsychiatry Unit, Policlinic of Bari; UFII=University of Naples Federico II, Child and Adolescent Neuropsychiatry Unit; SMF=Stella Maris Foundation, University of Pisa; UTV=University Tor Vergata; USS=University of Sassari, Child Neuropsychiatry Unit, Azienda Ospedaliero-Universitaria; UONPI-LO=Unita’ Operativa di Neuropsichiatria dell’ Infanzia e dell’ adolescenza, Lodi; ASD=Autism Spectrum Disorder, ADHD=Attention-Deficit/ Hyperactivity Disorder; ID=Intellectual Disability. See eTable 1 and Methods in eAppendix for details on data collection protocols.

### Data Selection

Data selection criteria for analyses were: 1) AFAR survey completion within the completion time interval for 90% of a given sample, to exclude outliers regarding COVID-19 infection rates and related responses for that sample; 2) age five years and above, as not all domains examined applied to younger children; 3) available AFAR variables included in clustering analyses.

### Analyses

#### Overview

Across the aggregate sample, exploratory (EFA) and confirmatory factor analyses (CFA) were used to identify the AFAR scores. For consistency with earlier literature and interpretation of results, repeated measures MANCOVA examined group level pandemic effects on symptom changes (*Prior* versus *Current*), accounting for site. Measures of central tendency (mean and proportions) describe the total number of services lost or continued within and outside school. Hierarchical clustering (HC) and Random Forest (RF) classification served to identify pandemic impact subgroups and their predictors, respectively. The code used for factor analyses, HC and RF can be found at github.com/ChildMindInstitute/CRISIS-AFAR-analyses.

#### Factor Analyses

EFA and CFA were conducted in split-half datasets, group-matched by contributing sample, sex, child age, Full Intelligence Quotient (FIQ), and primary diagnosis (i.e., ASD, attention-deficit/hyperactivity disorder [ADHD] without ASD, Other NDD). EFA was first conducted across items included in the survey domains. Then, items with resulting factor loadings ≥0.3 were included in subsequent CFA. Items were removed, as needed, to yield factors composed of more than one item, meeting at least two of four goodness-of-fit-criteria, and theoretical plausibility. For the four domains designed to assess *Prior* and *Current* behaviors, EFA and CFA were conducted using the *Prior* scores; then, their stability was assessed via a CFA on items using the *Current* scores across the whole sample, (see Methods in eAppendix).

#### Hierarchical Clustering

To identify homogeneous subgroups with distinct profiles across 11 features of symptom and service changes, we performed agglomerative hierarchical clustering.^37,38^ Seven features reflected changes in clinically relevant symptoms for ASD/NDD, indexed by differences between *Current* and *Prior* scores on domains identified in factors analyses (Supplementary Methods). The remaining four features included the total number of services that were either lost or continued within and outside school. All scores were converted to standard *z* scores prior to clustering. Clusters (i.e., subgroups) were characterized by deviations from the aggregate sample average. The optimal cluster solution was determined using the *NbClust*,^39^ according to a majority rule among multiple goodness of fit measures.

#### Random Forest

We assessed the relative contribution of 20 features in predicting the COVID-19 pandemic subgroup (Figure 2A, Supplementary methods). These included family and child socio-demographics, their pandemic experiences, COVID-19 pandemic factors such as government responses, new COVID-19 infection rates empirically derived from open-data sources,^40–42^ as well as pre-pandemic service received and clinical severity. The latter was indexed by a summary *Prior* (baseline) score computed across the 7 AFAR symptom domains, this summary score was significantly correlated with previously collected standardized measures in datasets with those data available (eFigure 6, Methods in eAppendix). Based on a previously established analytical RF framework,^43–45^ feature importance was calculated using the permutation importance method^45^ and indexed as the average out-of-bag-error (OOBE), computed across 4000 bootstrapped samples (⅔ training, ⅓ testing). The larger the OOBE value, the greater the importance for a given feature (Methods in eAppendix).

**Figure 2.**
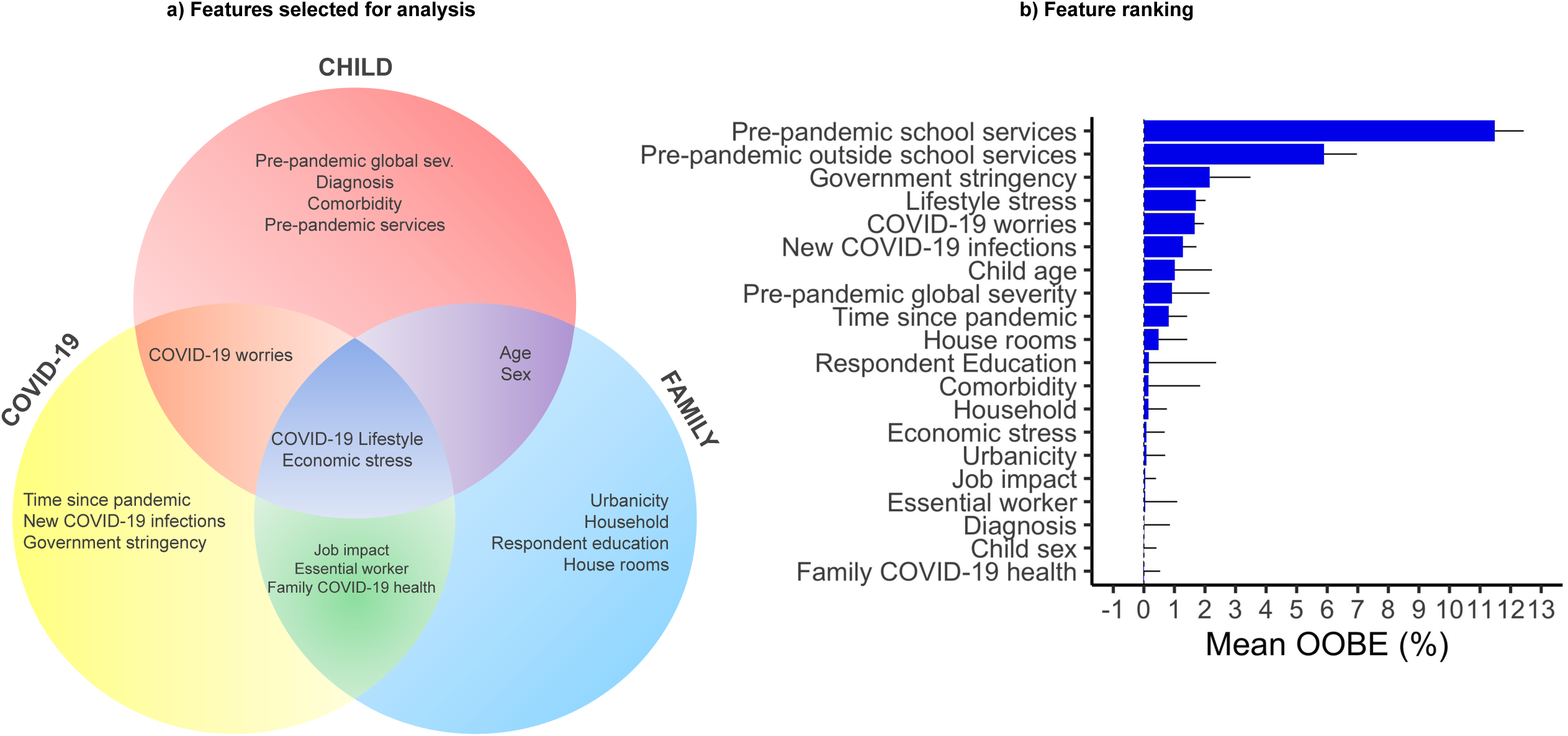
Prediction of COVID-19 Impact. a) The Venn diagram shows the 20 features selected for the Random Forest analysis across three partially overlapping domains: Child (socio– demographics, clinical characteristics before and during the pandemic; in red); COVID-19 pandemic factors (COVID-19 rates and related restrictions, and time since pandemic started; in yellow); and Family/Household factors (urbanicity, household composition and respondent education; blue). b) Feature ranking by mean OOBE importance plot across the 20 features included in the Random Forest model shown in descendent order. Abbreviations: OOBE, average out-of-bag error.

## RESULTS

### Aggregate Sample

Data from 1275 youth aggregated across contributing samples met inclusion criteria for subsequent analyses. Demographic, clinical, and other characteristics are in Figure 1B, Methods in eAppendix, eFigures 2 and 3, eTables 2-4. Most individuals (79%, n=1004), met diagnostic criteria for ASD; 17% (n=214) had ADHD without ASD, and 4% (n=57) had other NDDs, without ASD. Amongst 938 (80%) youth with available intelligence estimates, 62% (n=624) were in the Average/Above Average range, 14% (n=141) in the Borderline range, and 24% (n=173) had mild to profound intellectual disability. Over half (64%, n=811) of the caregivers had at least a college degree, the remaining had either a high school degree (30%), or elementary education (6%). Fifty-six percent of the aggregate dataset was of European/British ancestry (eTable4).

### AFAR Factors

EFA and primary CFA conducted on the *Prior* scores of split-half samples yielded a single Adaptive Living Skills factor, two RRB-related factors (High- and Lower-order),^33,46^ and four Co-occurring Problem Behavior factors (Anxiety, Oppositional Behavior, Sleep Problems, Activity/Attention (eTables5 and 6). As in the general population,^27^ COVID-19 Worries yielded a single factor, while Daily Behaviors and Life Changes yielded multiple factors. Results remained consistent in secondary CFA across the entire sample on *Current* ratings (eTable 6).

### Impact on Overall Sample

Only sleep problems reached statistically significant increases between *Prior* and *Current* scores (p=0.03, FDR corrected; Results in eAppendix). On average, children lost one service and had one other service continued, either at or outside school. Most continued services occurred as modified via telehealth/email (eFigure 4, eTable 7).

### Pandemic Impact Subgroups

The four-factor cluster solution of COVID-19 impact was the most optimal (Figure3, eTable7, eFigure4). Based on their profile of deviations from the aggregate’s average of symptoms and/or service changes, the subgroups were: *broad symptom worsening only* (20%); *primarily modified services* (23%), *primarily lost services (*6%), and *average symptom/service changes* (53%). The *broad symptom worsening only* subgroup was characterized by more severe scores across all symptom domains as indexed by z scores >.5 (>.5 SD from the average aggregate), and by marginal service changes (i.e., within .5 SD from the average). The three remaining subgroups, totaling n=1024 (80% of the aggregate) showed symptom changes within the aggregate’s average (z scores < .5) but differed in service access. One subgroup had most services modified, another had most services lost, and a third subgroup had the number of services lost and continued like those of the aggregate. This overall symptom/service impact pattern was confirmed by follow up one-way ANOVAs and Tukey pairwise group mean comparisons (FDR q<0.05). Results remained largely similar after covarying for contributing samples, and in secondary cluster analyses on data subsets distinct by survey attrition rates (eTable7, eFigures4 and 5, Results in eAppendix).

### Predictors

The RF model predicted subgroup membership with 81% accuracy (precision/sensitivity=82%, recall/specificity=75%). The top-ranked predictor (OOBE: 12 %) was the number of services received at school before the pandemic (Figure 2B). Six predictors followed with OOBE going from 6 to 1%. They included the number of services previously received outside school, COVID-19 rates, stringency index in the child’s location, lifestyle stress, COVID-19 Worries, and age (Figure 3, eTable 8). Baseline severity score prior to the pandemic was ranked eighth (<1%). Each subgroup was characterized by unique combinations of the pandemic, pre-pandemic, and demographic predictors (Figure4, eTable8).

**Figure 3.**
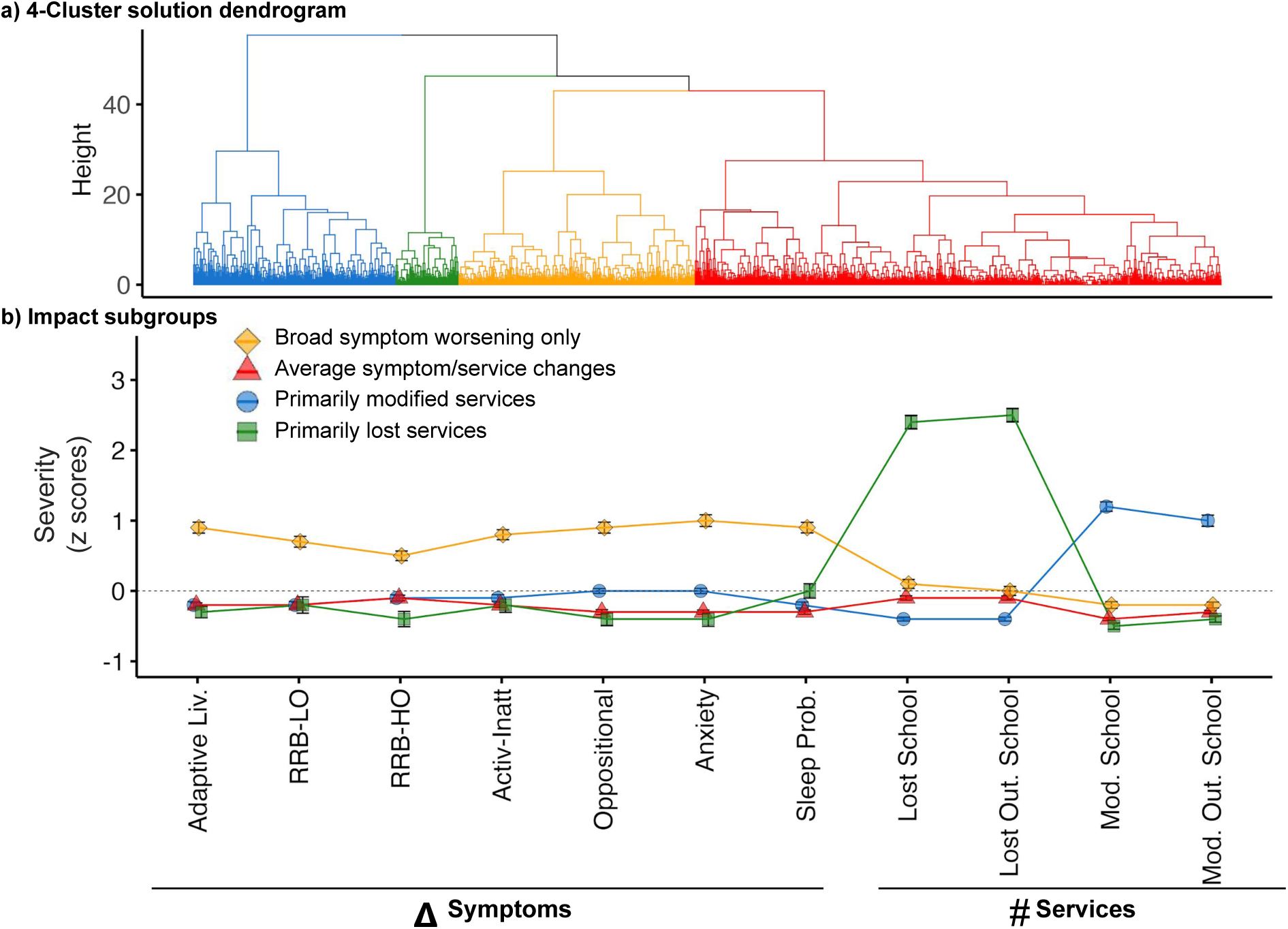
COVID-19 Impact Subgroups. a) The dendrogram shows the optimal 4-cluster solution which included: *broad symptom worsening only* (n=251, 20%; yellow), *primarily modified services* (n=293, 23%; blue), *primarily lost services (n=78*, 6%; green), and *average symptom/service changes* (n=653, 53%; red). b) Group means and standard error bars of the e z scored symptom factor changes (difference between Current and Prior scores; or Δ) and number of services lost or modified in and outside (Out.) of school are shown for each cluster (i.e., impact subgroup). The dotted gray horizontal line at a z score 0 represents the aggregate sample mean (N=1275) across the features examined. Abbreviations: Adaptive Liv., Adaptive Living skills; RRB-LO, Restricted and Repetitive Behaviors - Lower Order; RRB-HO, Restricted and Repetitive Behaviors - Higher Order; Activ-Inatt, Activity Inattention; Sleep Prob., Sleep Problems; Out., Outside; Mod., Modified.

**Figure 4.**
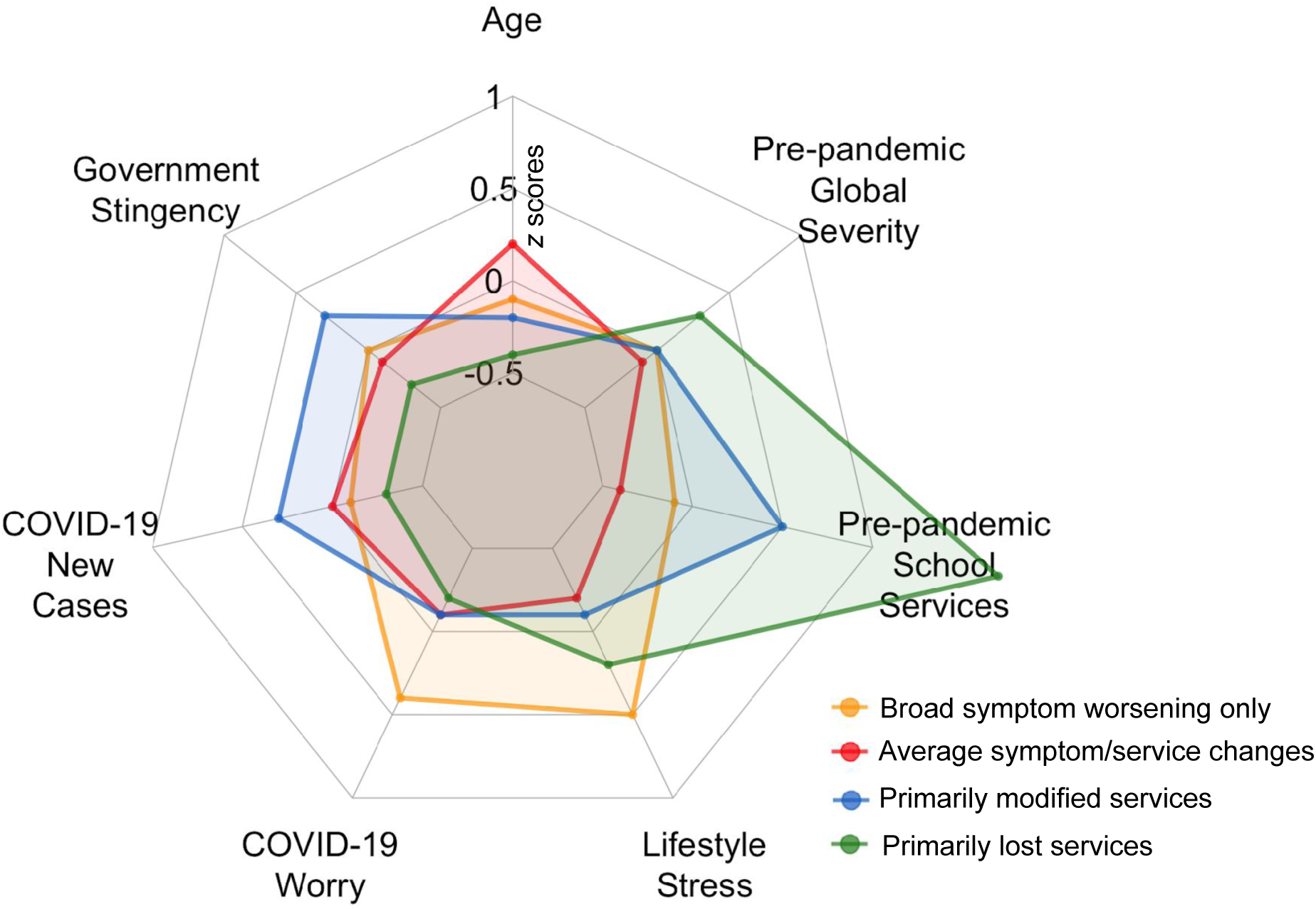
Top-ranked Random Forest predictors by Impact Subgroups. The radial plot shows the subgroup means across the eight top-ranked predictors resulting from the Random Forest analyses, color-coded by subgroup (Yellow**=***broad symptom worsening only*; Red=*average symptom/service changes*; Blue=*primarily modified services*; Green=*primarily lost services*). Data are shown as z scores.

## DISCUSSION

Prior disaster research in the general population, including COVID-19 pandemic studies,^27,47,48^ has consistently highlighted heterogeneity of mental health outcomes. The present study extends this insight into a multinational large sample of ASD/NDD youth by concomitantly assessing variability of changes in symptoms and therapeutic service access early in the pandemic. Across contributing samples, data-driven analyses identified four ASD/NDD subgroups: *broad symptom worsening only* (20%), *primarily modified services* (23%), *primarily lost services (*6%), and *average symptom/service changes* (53%). Their profiles revealed that symptom and service changes have distinct patterns of variation. The subgroup with notable clinical worsening did neither have the greatest number of services lost, nor the greatest number of continued services. Conversely, youth with relatively stable symptoms were parsed in three subgroups differing in service access. Recognizing these subgroups led to identify unique combinations of protective and risk factors, highlighting different pathways to either stable or worsening clinical presentations in ASD/NDD youth. The most robust predictors included factors common to the general population (e.g., COVID-19 worries), others ASD/NDD-specific (e.g., number of services received before), and age. Thus, the varying pandemic impact on symptoms in ASD/NDD youth is predicted by combinations of universal and ASD/NDD-related pre- and pandemic contexts in which service changes occur.

With one exception,^20^ the first wave of studies documenting the impact of the pandemic on ASD/NDD youth has consistently reported worsening behavioral and/or emotional problems,^12,16,17,49,50^ ASD symptoms,^49,51^ and/or sleep disruptions.^10,12,50,52^ Our results underscore that solely focusing on group-level effects leads to an incomplete picture of the COVID-19 pandemic impact on ASD/NDD. Multivariate analyses, across the whole sample, revealed that only sleep problems significantly worsened from pre- to pandemic times. While this group-level approach confirms earlier pandemic reports of increased sleep problems in ASD/NDD,^12,15,50^ it fails to recognize a more vulnerable subgroup. Cluster analysis revealed that 20% of the children worsened above and beyond their ASD/NDD peers. Worsening affected a broad range of symptoms, including sleep, externalizing, internalizing symptoms, RRB, and daily living skills. For the remaining participants, symptom changes pre- to pandemic were within the aggregate average - i.e., pandemic-related increases in sleep problems with other symptoms being relatively stable. Consistent with prior literature,^8,9,51^ our ASD/NDD sample experienced a variety of service disruptions, across settings, with either loss or telehealth modifications. However, variability was notable. Concomitantly clustering services and symptom changes further parsed the relatively clinically stable youth in three homogeneous subgroups differing by service access. This enabled a fine-grained identification of risk and resilience factors.

Employing AFAR in a multinational sample with varying COVID-19 rates and restrictions allowed the assessment of a broader range of predictors of impact than previously examined. The most relevant predictors of subgroup membership included risk factors common to the general population, and others more specific to ASD/NDD, such as pre-pandemic services, pandemic-related environment, and age. For example, elevated COVID-19 worries, and stress related to restrictions on leaving home and cancellations of important events distinguished the worsening subgroup. Consistent with prior disaster literature, ^25,53^ these factors have been identified as strong predictors of negative outcomes in recent general population pandemic studies.^27,28^ Our results underscore the impact of these stressors in clinical and non-clinical populations.

The present work also highlights the additional intricacies of understanding crisis impact on heterogeneous clinical populations. The number of services received prior to the pandemic was lower in the *broad symptom worsening only* subgroup versus two of the relatively stable ones: the *primarily modified* and the *primarily lost services* subgroups. In turn, these differed from each other for COVID-19 environmental context; the *primarily modified services subgroup* lived in areas with higher COVID-19 rates and greater restrictions than the *primarily lost services* one. Children experiencing the least pandemic-related changes relative to the aggregate sample, on average, were older (late childhood/preadolescence) than the other three subgroups. Especially in middle childhood, these findings indicate that more baseline services may foster resilience during a disaster. When living in areas with greater restrictions due to high COVID-19 rates, continuing services, even if modified, may lead to a relatively more stable clinical profile.

Pre-pandemic global clinical severity in ASD/NDD and other features related to clinical severity including diagnostic status, comorbidity rates, intellectual, and adaptive skills negligibly contributed to prediction. Most children across subgroups had equivalent baseline symptom severity, average intelligence, and mildly impaired adaptive functioning except for the *mostly lost services* subgroup which was characterized by greater impairment. These findings are in contrast with earlier ASD studies^10,51^ which suggest that pre-existing behavioral challenges related to greater problem behaviors following the pandemic. However, unlike the present effort, prior work focused on samples from relatively narrow geographical areas with largely homogeneous COVID-19 rates and institutional responses. The range of impacts and experiences of the COVID-19 pandemic, across our aggregate sample, allowed us to paint a more comprehensive picture of risk and resilience across ASD/NDD youth.

Our study has several limitations. First, considering time constraints on questionnaire completion, albeit comprehensive, AFAR could not assess all domains of impact and/or prediction. Symptoms least expected to change over a short period of time, were given lower priority, most notably, social-communication impairments.^54^ Given the protracted nature of the pandemic, future studies should include long-term assessment of social-communication skills. Similarly, although family demographics, parent education, and parent being an essential worker were included in our predictive model, parent’s mental health, recently reported to relate to children’s outcome during the pandemic,^55–58^ was not assessed. Second, although the aggregate sample includes youths with clinician-based diagnoses, previously collected measures of severity varied by contributing sample, and assessments of *prior* severity were based on parent responses in the AFAR survey. Nevertheless, we found that the AFAR baseline global severity scores correlated with standardized measures, when available. Third, our study did not include ASD/NDD preschoolers. Thus, although consistent with a prior report, ^51^ our results indicate that those in middle-childhood are at greater risk of impact; future studies specifically designed to target younger ages are needed. Finally, the present study focused on impact over the first six months of the pandemic using a cross-sectional design. Longitudinal coordinated study designs and infrastructures (e.g., common assessment measures) are needed to capture long-term outcomes and define stability of subgroups over time.

In sum, as in the general population, the COVID-19 pandemic impact varies across ASD/NDD youth. Risk and resilience are rooted in the pre- and pandemic contexts in which service disruptions occur. Provision of mental healthcare in preparation for, and during disasters are critical for ASD/NDD youth - further motivating efforts assessing effectiveness for telehealth and/or hybrid treatment programs. Finally, this study highlights the value of international data sharing and collaborations to address the needs of those most vulnerable.

## Supporting information

CRISIS AFAR Supplemental Materials

## Data Availability

All data produced in the present study are available upon reasonable request to the authors

## Additional Contributions

We are grateful to the research and clinical staff members at each contributing site supporting different aspects of collection of the AFAR surveys and the pre-pandemic clinical characterization. We thank Irene Droney at CMI for support in the development and curation of the AFAR website, RedCap data structure, Dr. Greg Kiar at CMI for helpful discussion about random forest classification, and Dr. Marco Pagani for helpful discussions on the use of NbClust in R. We are also thankful to Dr. Shafali Jeste for her sharing the preprint version of the service survey developed during the pandemic for individuals with syndromic intellectual disabilities and their caregivers that provided the model on which we developed the AAR service-related questions. Most importantly, we are immensely grateful to all parents/caregivers and their children who generously contributed their time during the pandemic, one of the most challenging times for humankind in this century.

