## Supplementary material for "CRISIS AFAR: An International Collaborative Study of the Impact of the COVID-19 Pandemic on Youth with Autism and Neurodevelopmental Conditions": CRISIS AFAR Supplemental Materials

**Supplementary Online Content**

**eAppendix.** Supplemental Appendix (Methods, Results)

**eTable 1.** Data collection and associated pre-pandemic phenotyping protocols for the 15 contributing samples.

**eTable 2.** Clinical characteristics of the aggregate dataset and 15 contributing samples.

**eTable 3.** Participant characteristics of the aggregate dataset and 15 contributing samples.

**eTable 4.** Ancestry of the aggregate dataset and 15 contributing samples.

**eTable 5.** Factor loadings by domain examined.

**eTable 6.** Goodness-of-fit for exploratory and confirmatory factor analyses.

**eTable 7.** Group means of symptom changes, number of services lost and modified in the aggregate dataset and by COVID19 impact subgroups.

**eTable 8.** Group means of the top-ranked predictors identified via random forest analyses in the aggregate dataset and by subgroups.

**eFigure 1.** CRISIS AFAR parent baseline survey v0.5.1 (3-21 years): domain structure and items.

**eFigure 2.** Data collection and selection flow.

**eFigure 3.** CRISIS AFAR survey background and COVID-related information for the aggregate dataset.

**eFigure 4.** Symptom and service access change.

**eFigure 5.** Subgroup distribution by contributing samples.

**eFigure 6**. Correlations between pre-pandemic standardized severity measures and AFAR baseline global severity score.

**eAppendix. Supplemental Appendix**

**Methods**

**Adaptation process to yield AFAR**

***Overview***. The Coronavirus Health and Impact Survey Initiative (CRISIS)^1^ adaptation for autism and related neurodevelopmental conditions (AFAR) was led by a working group of psychologists and psychiatrists with clinical and research expertise in autism spectrum disorder (ASD) and other neurodevelopmental disorders (NDD) (A.D M., L.G., S.G., P.P., A.T., B.V.). Their goal was to preserve the main structure of CRISIS^1^ developed for the general population while adding domains most relevant to ASD/NDD. To this end, each member of the working adaptation group independently reviewed and coded each item of the CRISIS Parent/Caregiver Baseline 5-21 survey to be retained or removed and noted rewording suggestions. Second, the working group agreed on the assessment domains to be newly included. The details of retained, newly included, and removed and rewarded content are summarized below. Then, the working group, in consultation with another AFAR network member (S.H.K), assessed which items were theoretically applicable to children younger than five years. Later, minor rewording for some items based on feedback and agreement with the larger AFAR network, followed. The final version 0.5.1 of AFAR Baseline Parent/Caregiver (3-21 years) consists of 96 total independent items, including 34 questions asked twice for *Prior* (i.e., three months prior to the start of the pandemic in the respondent’s geographical area) and *Current* time points (i.e., last two weeks). The 96 total items include 26 multiple choices, 60 Likert scale, four yes/no, four ‘fill-in-the-blank’, and two open ended questions. AFAR is licensed on Creative Commons (CC) BY4.0 and available at <http://www.crisissurvey.org/crisis-afar/>. Additional details on the adaptation process are provided below.

***Retained/removed content*.** Items with more than 90% agreement for retention or removal across the working group raters were modified accordingly. For the other items, the working group met to reach a consensus for retention or removal. All rewording’ suggestions on retained items were also discussed to reach a consensus. Overall, two CRISIS domains (Mood States and Substance Use), along with one item from the CRISIS Background and eight items from the CRISI Life Changes domain were removed. These items/domains were removed to favor questions focusing on observable behaviors to limit parent/caregiver interpretations on internal states and the overall survey length after the addition of new ASD/NDD relevant domains (see below). Rewordings largely aimed to account for potential verbal communication differences across individuals with ASD/NDD. Several rewordings and minor updates were further applied with the input and consensus of the larger AFAR network at later versions of the AFAR survey. As a result of this process, a total of 23 CRISIS questions or response options were reworded (five in the Background, two in the COVID-19 Health/Exposure Status, five in the Life Changes, 11 in Daily Behaviors and Media; see eFigure1 for a summary of the final CRISIS AFAR survey structure).

***Newly included domains.*** The working group agreed to include three new domains as relevant for ASD/NDD based on one prior disaster report^2^ available at the time of the CRISIS-AFAR development (April 2020) and the larger clinical literature.^3–7^ The new domains were Adaptive Living Skills, Restricted and Repetitive Interests and Behaviors (RRB), and Co-Occurring Psychopathology. Two sub teams of the working group drafted specific questions for the targeted domains which were then reviewed, agreed, and further edited in following videoconferences with all members of the team. This process resulted in: 1) an Adaptive Living Skills domain of four items asking the degree of one child’s independence in playing, structuring their activities, organizing mealtimes, and participating in hygiene and daily living routines; 2) an RRB domain of six items capturing the frequency of lower- and higher-order RRB^8,9^ described in the DSM-5 diagnostic criteria^10^ (i.e. two items of sensorimotor RRB and four items assessing aspects of insistence on sameness). Lastly, to evaluate co-occurring psychopathology, caregivers were asked to indicate if specific externalizing and internalizing symptoms, often reported in children and adolescents with ASD/NDD,^3,5,11^ were present. If so, parents/caregivers were asked to indicate the symptom level of severity. Additionally, given the high prevalence of sleep problems in ASD,^12^ two questions on insomnia were added to the CRISIS sleep items included in the Daily Behavior and Media domain. Consistent with the CRISIS structure, these symptom domains were assessed at two time points: 1) over the three months prior to the COVID-19 pandemic beginning in the respondent's geographical area, and 2) over the past two weeks at time of data collection (hereafter referred to as *Prior* and *Current* time points, respectively). In order to evaluate changes in therapeutic services, the working group adapted and added to selected items of the Caring through COVID survey developed for individuals with syndromic intellectual disabilities.^13^ Specifically, caregivers were asked whether therapeutic services typically received at and outside school settings (e.g., speech language, physical and/or occupational therapy, social skills interventions) were lost or continued. If they were continued, caregivers were asked to specify if they had been received virtually (telehealth/email) or not and how helpful they were perceived by the caregivers. Caregivers were additionally asked about a range of medical services (e.g., gastroenterology needed and accessed), as well as if psychotropic medication prescription/monitoring were needed.

***Age range applicability*.** As shown in eFigure 1, all questions in the Co-occurring Problems domain, and two items of the Daily Behaviors and Media domain (i.e., interacting using technology to engage with peers and family) were deemed not applicable for children aged below five years. The group agreed that the remaining AFAR questions were developmentally appropriate for assessing children aged three years and above.

**Exploratory and confirmatory factor analyses (EFA, CFA)**

For factor analysis, we used Lavaan Package^14^ in R version 4.0.0^15^. The R code used for analyses can be found at github.com/ChildMindInstitute/CRISIS-AFAR-analyses. Exploratory Factor Analyses (EFA) were conducted across all items included in the theoretically developed assessment domain. Then, items with factor loadings ≥ 0.3 in the EFA were included in subsequent confirmatory factor analyses (CFA). Other items were removed as needed, to reach a final factor structure composed of more than one item meeting at least two of four goodness-of-fit criteria, and theoretical plausibility. Goodness-of-fit criteria commonly used in the literature included χ2 significance (non-significant values suggest good of fit^16^), root-mean-square error of approximation (RMSEA; cutoffs of .01, .05, and .08 indicate excellent, good, and acceptable fit, respectively^17^); the Tucker-Lewis index^18^ (TLI; ≥.95 indicates good fitting^19^), and the Bentler’s comparative fit index^20^ (CFI; ≥.96 indicates goodness of fit). As shown in eTable 5, following EFA three items were excluded due to factor loading <0.3; the item “*deliberately injuring self*” was removed from the attention and activity factor for interpretability. Two more items were not included in CFA as they resulted as single item factors in EFA; one was “*worry about own mental health*” in the COVID-19 Worry domain, the other was “*positive changes,*” in the Life Change domain.

**AFAR data collection and selection**

***Sites/contributing samples.*** As shown in Figure1a and eTable1, data were collected in 14 clinical and/or research institutions across Europe and North America yielding 15 independent datasets, referred to as contributing samples. These included one sample collected across five centers in Ontario, Canada as part of a COVID-19 multi-network collaboration in pediatrics that included the Province of Ontario Neurodevelopmental Disorders (POND-CMH) Network,^21–23^ five samples collected in four research and clinical institutions across the United States of America (New York, California, and Missouri), and seven sample collected across Europe (Greece, Italy, and Ireland).

***Data collection protocols.*** For each sample, AFAR data collection protocols are summarized in eTable1. Briefly, caregivers of children aged between 3-21 years who had previously received a clinician-based DSM-IV/5^24,25^ or ICD-10^26^ diagnosis of ASD and/or other NDD were invited via email and/or phone contacts to complete the AFAR survey. Initial contacts for this study included families who enrolled in ongoing research studies (3 contributing samples), clinical services (8 samples), or both (4 samples). Across all sites caregivers were invited to complete the survey regardless of their child’s biological sex, IQ, parent-reported race, ethnicity, or socioeconomic status. For all but one sample, surveys were administered exclusively online, and at least weekly reminders were used. No reward at completion was given for all except three contributing samples that provided $10 and $25 gift cards at completion (at Child Mind Institute and University of California, San Francisco, respectively). Data collection time and duration varied by sample over the period between April 24 and October 20, 2020 (Figure 1, eTable 1). Reflecting geographical variation across the collection sites, AFAR was completed under different COVID-19 related restrictions ranging from shelter-in-place ordered uniformly in each geographical area, to less stringent and geographically heterogenous restrictions. This allowed us to naturalistically explore the impact of distinct restrictions on the NDD/ASD impact subgroups identified with hierarchical clustering (see below on the Random Forest (RF) section, and Figure 2A for the feature selected for prediction). Data were organized in a common data template.

**Prior diagnostic and clinical protocols**

To provide a more accurate clinical characterization of the aggregate dataset than otherwise feasible in large scale online survey efforts, the AFAR network aimed to collect data of children previously well characterized via systematic clinical assessment protocols conducted by clinicians prior to the COVID-19 pandemic. Although the diagnostic clinical protocols varied by contributing sample, within each site, clinician-based DSM-IV/5 or ICD-10 estimate diagnoses were previously reached based on parent interviews, direct child observations, and cognitive testing, review of parent questionnaires and other available records followed by group case conferences to reach best-estimate diagnoses following a review of observations and discussion of clinical impressions. Specifically, as summarized in eTable1, the gold-standard Autism Diagnostic Interview-Revised (ADI-R)^27^ and/or Autism Diagnostic Observation Schedule (ADOS-G and/or second editions)^28,29^ were systematically used in 12 of the 15 contributing samples and in subsets of children for the remaining two samples (Healthy Brain Network [HBN], Trinity College - Dublin [TCD]). In six samples, the Kiddie Schedule for Affective Disorders and Schizophrenia-Present and Lifetime (K-SADS-PL)^30^ or unstructured psychiatric interviews were used to aid diagnosis and for further clinical characterization. Adaptive functioning was systematically assessed via standardized interviews (e.g., Vineland-3 Comprehensive Parent Interview, Third Edition [VABS-3]^31^ or Adaptive Behavior Assessment Scale, Third Edition [ABAS-III]^32^) in all but two samples (TCD, University of Athens [UAth]). Standardized cognitive testing was administered to all children, as feasible, with the specific tests selected depending on the child’s age and developmental level. The clinical characteristics of each contributing sample and of the aggregate dataset are summarized in Figure 1B and eTable 2.

**Harmonization of shared phenotypic information**

***Overview.*** Along with diagnostic labels (available for 100% of the aggregate dataset), whenever feasible, information on intellectual functioning and other quantitative metrics of symptom severity were also shared. These included data for intelligence (80% of the aggregate sample), a measure of expressive language derived from the ADOS-2^33^ (57%), psychopathology indexed by the parent-based Child Behavior Checklist (CBCL)^34^ summary T scores (59%), and adaptive functioning (53%). Below, we describe the curation and harmonization approach used for comparability across samples.

***Diagnostics.*** Per study design, all contributing samples provided clinician-based diagnostic labels. They were consistent with the DSM-5 diagnostic criteria for all but two sites: Lodi that used DSM-IV, and TCD that used ICD-10 for a subset of children. Diagnostic labels were mapped into the DSM-5 nomenclature (e.g., Asperger was remapped into ASD). When one child had multiple diagnostic labels, the most frequent diagnosis in the aggregate sample was considered the child’s primary diagnosis and the other diagnostic labels were accounted for as comorbidities (e.g., ASD with or without attention-deficit/hyperactivity disorder [ADHD] and/or anxiety). When a child did not have the most frequent diagnostic label, the second most frequent label in the aggregate sample was considered the child’s primary diagnosis - if present (e.g., ADHD without ASD - ADHD_w/oASD_) and other labels, if assigned to that child, were considered comorbidities (e.g., ADHD_w/oASD_ with comorbid anxiety and/or learning disorder). This process was repeated iteratively for the following most frequent diagnostic labels (e.g., intellectual disability without ASD and without ADHD). See Figure 1 and eTable 2.

In the aggregate dataset, ASD (with or without comorbid diagnoses) was the most frequent diagnostic label (n=1004, 79%). Of the remaining 271 children without ASD, the most frequent diagnostic criteria were ADHD (n=214, 80%%), for intellectual disability (ID; n=15, 6%), anxiety disorders (n=10, 4%), obsessive compulsive disorder (n=9, 3%), disruptive-impulse control and genetic conditions (n=7, 3% for each diagnostic label), language/learning or other neurodevelopmental disorders (LD/NDD; n=5, 2%), depressive disorders (n=2, 1%), and n=1 each for tic and avoidant personality disorders. Among the children with ASD, 434 (43%) were reported to meet criteria for at least one co-occurring diagnosis. Of those with co-occurring diagnoses, 270 (62%) met criteria for ADHD, 116 (26%) for ID, 81 (19%) for an anxiety disorder, 77 (18%) for LD/NDD. Among those with ADHD_w/oASD_ , most were without comorbidity (n=171, 80%). Similarly, among those with other diagnoses without neither ASD nor ADHD (n=57), the majority had no comorbid diagnoses (n=44, 77%). See eTable 2 for diagnostic and other clinical information on the aggregate dataset and each contributing sample.

***Intelligence.*** Information about intellectual functioning was available for 80% (n=1014) of the aggregate dataset; it included either quantitative metrics or qualitative clinician-based categories of intelligence (n=962 and 47 children for n=15 and n=6 samples, respectively). For comparability across samples, since full intelligence quotient (FIQ) was the most represented metric in the aggregate dataset (60%, n=759 children), we focused on mapping quantitative variables of full-scale intelligence in a comparable distribution of Mean=100 and Standard Deviation=15. When only VIQ and NVIQ scores were provided (1% of the aggregate, n=15 children, 3 Samples), their average was used to estimate FIQ. When quantitative metrics of development (e.g., obtained with Mullen Scales of Early Learning^35^) were provided (5% of the dataset, n=63 children, 8 samples), they were converted to Developmental Quotients (DQ), as in prior literature in NDD/ASD.^7-36^ For those samples with less than 15% FIQ data missing, within each sample, we imputed missing data using the average FIQ computed across children with non-missing data group-matched by age and diagnosis (n=54 children, 4%, n=6 Samples).

Taken together, this process resulted in a total of n=891 (70% of the aggregate) children with a quantitative FIQ (or equivalent) available and n=47 (4% of the aggregate) with a qualitative clinician-based rating of intellectual functioning. To harmonize qualitative and quantitative intelligence estimates of full-scale intelligence, we mapped all quantitative scores onto qualitative categories as described above. Then, to harmonize quantitative and qualitative estimates of full-scale intelligence, FIQ scores were mapped onto qualitative categories ranging from *above average* to *profound intellectual functioning*. Specifically, children with FIQ scores within or above one standard deviation of the standard mean were labeled with Average (FIQ=85-114); or Above Average intelligence (FIQ >115). Those with FIQ estimates below 85 were categorized as Borderline intelligence (84-71), Mild (51-70), Moderate (FIQ:35-50), Severe (FIQ: 21-34), and Profound intellectual disability (FIQ<20).^24^ As a result of the harmonization step, the total number of children with FIQ estimates was n=938 (74% of the aggregate) across all 15 contributing samples (Figure 1). Of the remaining n=337 (26%) without any FIQ equivalent available, n=117 children had NVIQ scores (9%, 7 samples) and n=220 had no available quantitative FIQ, NVIQ, or VIQ scores available. NVIQ scores were not converted to qualitative full-scale estimates, but they are described in Supplementary Table 2.

***Expressive language.*** Measures of language were not uniformly administered, nor shared across all samples. Thus, to obtain a comparable characterization of verbal skills, we leveraged previously administered ADOS to compute a measure of expressive language derived by one ADOS item that has been validated in prior work^33^ (i.e., ADOS-EL). This was computed and shared for 57% of the aggregate dataset (n=730 children, 13 samples). As a note, ADOS calibrated severity scores^37^ were shared only for 38% of the aggregate (n=479 children; 7 samples). As such, we did not use them in targeted analyses but, considering their common use in ASD studies, we report their descriptive statistics in eTable2.

***Adaptive functioning.*** Quantitative indices of adaptive functioning were shared for nine contributing samples yielding data for 53% of the aggregate dataset (n= 674). Different editions of the VABS^31,38^ were used across six samples, the remaining three samples used the ABAS-3. Both instruments assess functioning across multiple living domains and provide a summary score, Adaptive Behavior Composite and General Adaptive Composite, respectively, in a standard scale with M=100 and SD=1). Thus, for comparability across samples, composite scores were used to characterize global adaptive functioning.

**Impact on Overall Sample**

Although the main goal of the present study was to assess whether different COVID-19 pandemic impact subgroups exist in ASD/NDD, for interpretation of results and consistency with the pandemic literature more broadly, we examined symptom and service access changes across the aggregate sample (eTable7, eFigure 4). Accordingly, to explore symptom changes between *Prior* and *Current* scores for the seven symptom domains analyzed in clustering analysis, we conducted a one-way repeated measures MANCOVA using time as within-subject factor (*Prior* versus *Current)*, including contributing sample as covariate. Follow-up Post Hoc univariate one-way repeated measure ANCOVAs were performed to compare the two Prior vs. Current timepoints within each symptom domain. All tests were corrected with False Discovery Rate (FDR) at q<0.05. To characterize service changes across the aggregate sample, we used measures of central tendency (e.g., mean for continuous variables, proportions for categorical variables) for the number of services whose access was lost and/or continued at school and outside school, separately.

**Hierarchical Clustering (HC)**

To examine whether homogeneous subgroups of ASD/NDD children with distinct profiles of change in symptom and/or service exist, we performed agglomerative hierarchical clustering (*hclust* function from the *cluster* package in R version 3.6.1).^39^ We used Euclidean distance (*dist* R function) and Ward's minimum variance method (*ward.D2* option; Murtagh and Legendre’s criterion).^40^ Clustering was conducted across 11 features of impact based on AFAR. Seven of the features were comprised of change scores indexed by difference scores between *Current* (“last 2 weeks”) and *Prior* (“three months prior to COVID-19”) time points in the seven domains identified by factor analyses (i.e., Adaptive Living Skills, High- and Low-order RRB, Anxiety, Oppositional Behavior, Sleep Problems, Activity/Attention). The remaining four features included the total number of therapeutic services that were lost or continued in and outside school following the pandemic. All scores were converted to standard z scores prior to clustering. The *NbClust* package^41^, a consensus-based algorithm that ranks solutions based on the degree of agreement across 30 different established clustering-quality indices (e.g. Davies–Bouldin or Gap statistics) was used to determine the optimal number of clusters between 2 and 15 solutions according to a majority rule. The implementation and R code used for the analysis can be found at github.com/ChildMindInstitute/CRISIS-AFAR-analyses.

**Random forest (RF) feature selection and model**

To assess the relative importance of a set of COVID-19 pandemic-related feature, socio-demographic and child clinical variables in predicting the COVID-19 impact subgroups, we used RF implemented in scikit-learn^42^ (see github.com/ChildMindInstitute/CRISIS-AFAR-analyses).

***RF analytical framework.*** Consistent with prior work,^43^ we assessed each feature importance across 4000 bootstrap samples that were drawn with replacement from the children in the aggregate dataset with the variables included in the RF available (N=1244; see eFigure 2 in Supplement). Each bootstrap sample was split in a training (2/3) and a testing (1/3) set. For each sample, a classification model for four subgroups was first developed by growing 300 trees in the training set and then applied to the test set. Feature importance was calculated using the permutation importance method ^44^ similarly to prior work.^43,45^ Briefly, within each sample, we first recorded a baseline accuracy score for the trained model, permuted the values of each feature, then passed the test sample back through the RF and recomputed accuracy. The importance of a given feature was indexed by the difference between the baseline and the new accuracy value obtained across permutations; this is known as the ‘out of bag error’ (OOBE). An average OOBE value was calculated for each of the 20 analyzed features. The larger the OOBE value, the more important the feature is. All features were ranked accordingly in decreasing order of importance.

***RF feature selection.*** As shown in Figure 2A, the RF model assessed 20 features indexing family and child socio-demographics, their pandemic experience, pre-pandemic child clinical characteristics including services previously received, as well as different COVID-19 pandemic context such as government response, new infection rates, and time since the start of the pandemic. Most of these features were largely derived from AFAR responses except for the COVID-19 rates at the time of completion in the respondent’s geographical area and the Government stringency index (see below) that were derived from open data sources (see below).

The child/family socio-demographics, pandemic impact on jobs and health, child’s COVID-19 worries, and perceived economic and life stress were based on AFAR factors identified in EFA/CFA.

and other responses on the AFAR background section. For the pre-pandemic clinical presentations, we computed a baseline (i.e., 3 months prior to the pandemic) global severity score across the seven clinical factors of AFAR (indexed as an average z score). We also used primary diagnosis of ASD versus non-ASD and number of comorbid diagnoses. For this primary RF model, we did not include standardized clinical measures collected prior to the pandemic described above (i.e., CBCL, VABS/ABAS, ADOS-EL) as they were available for less than 70% of the aggregate individually and when combined only for n=453 individuals (36% of the aggregate). However, as shown in eFigure 6, these clinical measures were statistically significantly correlated with the Baseline Global Severity from AFAR. Nevertheless, we repeated RF models including these standardized variables in the smaller sample with all variables examined and the pattern of results remained the same (data not shown).

To quantify government responses to the covid-19 pandemic, for each child at the time of the AFAR data collection in their geographical area, we used the government stringency (GS) index computed by the Oxford’s Coronavirus Government Response Tracker^46^ (latest download on 25 August 2021). The GS index combines metrics of infection containment and public information campaigns, it ranges from 0 to 100, higher scores reflect stricter government policies, and it is provided by day in each territory. For each child, we selected the GS index computed at the same date of the AFAR completion in the respondent’s living area. GS were available at the subnational level for USA and Canada (e.g., New York, California, Missouri, and Ontario) and at the nation level for Greece, Italy, and Ireland.

To quantify new infection rates in a given child’s geographical area at the time of AFAR data collection, we used the publicly available Our World in Data's (OWID) [COVID-19 tracker](https://ourworldindata.org/coronavirus).^47^ Data on new COVID-19 infected cases were available on a daily basis for the United States and Canada at a sub-national level. Since this data source only had country-level information across Europe, for the European samples we opted to use the COVID-19 European regional tracker^48^ which provides daily new infection rates at the regional level. Based on each European contributing sample’s catchment areas, we retrieved location-specific information. For example, for children in TCD we used the Eastern-Midland region of Dublin. Regarding the sample collected by UAth, since over the time interval during which AFAR data were collected in the Attica region, major data gaps existed, we used country-level infection rate data available in the OWID that were available for the same period.

**Results**

**AFAR data collected**

As shown in eFigure2, of the total of 4458 surveys known to be sent to families, 1595 (36%) datasets were returned; success rate varied by contributing sample (13% to 84%; median=40% SD=22%). Six contributing samples had a return rate >50%; they included sites located in the USA, Canada, and Italy, and totaled 724 individuals. Nine contributing samples had return rates <50%; they included sites located in the USA, Italy, Greece, and Ireland; they totaled 551 individuals. As indicated below, secondary hierarchical clustering conducted on contributing sample sets defined by differences in return rates (> or < 50%) showed similar patterns as those observed in primary analyses.

**Impact on the Overall Sample**

***Symptom changes*.** One-way repeated measures MANCOVA across the aggregate sample revealed a statistically significant within subject difference (F_(7,2548)_= 77.4.0, FDR corrected p <0.001) with a main effect of time (*Prior* versus *Current)* across the symptom domains. Post Hoc univariate one-way repeated measure ANCOVAs assessing the contribution of specific symptom domains revealed that only sleep problems reached statistical significance (FDR *p* corrected=0.03; M±SD: *Prior*, 4.9±2.1, *Current* 5.2±2.3, *Current-Prior*, 0.2±1.5).

***Service changes***. Across the aggregate sample, of those who previously reported receiving services in school (n=1008, 80%) and outside school (n=939, 74%), the average number of services lost and continued was M±SD: Lost=1.2±1.7, Continued =1.1±1.4; at school and M±SD: Lost=1.2±2.0, Continued =0.8±1.3 outside school. Among those who previously received services, 61% (n=610) and 64% (n=603) lost at least one service within and outside school, respectively. Notably, 37% of the children (n=370) in school, and 42% (n=397) outside of school, lost all their services. Of note, most of the services continued were modified virtually via telehealth/email services (88% in school and 69% outside of school).

**Post Hoc analyses**

***Subgroups comparisons relative to AFAR symptom changes****.* One-way MANOVA analyses revealed a significant main effect of subgroups across the seven symptom domains examined (F_(3,1271)_=37.0, p<0.001, Pillai’s Trace=0.51, eta square=0.17). Post Hoc one-way ANOVAs, followed by Tukey Pairwise group mean comparisons assessed differences across symptom change domains by subgroups (FDR correction q<0.05; eTable 7). Specifically, the *broad symptom worsening only* subgroup showed significantly higher positive difference scores (i.e., worsening) in all domains relative to the other subgroups. The remaining three subgroups did not statistically differ from each other in symptom domain except for the Anxiety and Oppositional factor scores which worsened in the *primarily modified services*, relative to the *average symptom/service changes* and *primarily lost services* subgroups (eTable 7). Given potential differences between contributing samples, to examine if the pattern of subgroup differences was confounded by contributing sample, we conducted a one-way MANCOVA including contributing sample as covariates (dummy variable 1 to 15). The pattern of subgroup differences remained unchanged (F_(3,1257)_=36.8, p <0.001, Pillai’s Trace=.51, eta square=0.17) and no statistically significant main effect of contributing sample (p=0.2) on symptom change scores was detected.

***Subgroups comparisons relative to AFAR service changes.*** Following a statistically significant one-way MANOVA the average number of services lost or continued at each setting- e.g., at school and outside school; (F_(3,1271)_= 149.5, p<0.001, Pillai’s Trace= .96, eta square=0.32), Post Hoc one-way ANOVAs, followed by Tukey Pairwise group mean comparisons assessed service changes (FDR correction p<0.05; eTable 7). Regarding services lost, the *primarily lost services* subgroup had the greatest number of services lost (M±SD across both settings: 5.7±1.6) which was significantly higher relative to all other Subgroups. The *primarily modified* *services* subgroup had the lowest number of services lost (M±SD across all settings: 0.5±0.9). This was statistically different relative to both the *broad symptom worsening only* and *average symptom/service changes* subgroups (M±SD: 1.0±1.4 and 1.4±1.8, respectively), which, in turn, did not statistically differ from each other. For modified services, the *primarily modified services* subgroup had the highest and statistically different number of modified services (M±SD across all settings: 3±1) relative to the other three subgroups, which, in turn, did not statistically differ from each other (M±SD range: 0.3-0.7 ± 0.6-0.8). Given potential differences between contributing samples, to examine if the pattern of service differences was confounded by the contributing sample, a one-way MANCOVA including sample as a covariate (dummy variable 1 to 15) was repeated. The pattern of difference remained significant (F_(3,1257_)=157.3, p <0.001, Pillai’s Trace=1, eta square=0.3). The effect of the contributing sample was also statistically significant (p< 0.001); as such we followed up with ANCOVA comparisons; the pattern of results remained largely consistent to what reported above (data not shown).

**Hierarchical Clustering (HC) follow-up analyses**

To assess that the primary results were not affected by differences seen in return rates across contributing samples (Figure2), we repeated these clustering analyses separately on subsets of data derived across contributing samples with return rates above or below 50%. Specifically, n=724 (57% of the aggregate sample) data were included in the six contributing samples with return rates > 50% while n=551 (43% of the aggregate) were derived from the nine contributing samples with return rates <50%. For each subsample, the pattern of results remained virtually unchanged from that obtained in primary HC analyses (data not shown).

**eTable 1: Data collection and associated pre-pandemic phenotyping protocols for the 15 contributing samples.**

|  | **Contributing Sample Label** | **Contact Source** | **AFAR survey version/ administration mode** | **Catchment area** | **Data collection interval (d/m/2020)^a^** | **COVID 19 response at the time of collection** | **Target age range (years)** | **Parent Interviews** | **ADOS** | **Child Cognitive Testing** |
| --- | --- | --- | --- | --- | --- | --- | --- | --- | --- | --- |
| **USA, New York State** | CADB | C, R | E 0.5.1; ReDCap | Tri-state New York Metropolitan area | 06/18-08/15 | 03/28-06/07: Shelter-in-place order for 100% of non-essential workforce;  06/08 - 07/19: Gradual reopening from phase 1- phase 4 (e.g., malls, zoos, and gardens allowed to reopen, indoor dining at limited capacities) | 3-18 | ADI-R, VABS-II/3 | y | Bayley; DAS-II, MSEL; WASI; Ravens |
|  | CMI-AC | R | E 0.4.0, 0.5.1^b^; ReDCap |  | 04/24-06/02^a^ |  | 3-12 | ADI-R (<5 years); ASI and KSADS-PL (>5 years), VABS-II/3 | y | DAS-II, MSEL |
|  | CMI-HBN | R | E 0.5.1; ReDCap |  | 05/21-07/01 |  | 5-21 | KSADS-PL; ADI-R^c^, VABS-II | subset only | KBIT-2; WISC-V; WAIS; WASI |
| **USA, California** | UCSF | C | E 0.4.0; ReDCap | San Francisco Bay area | 06/12-10/19^a^ | 03/16-05/14: Shelter-in-place order for 100% of non-essential workforce;  05/15: Gradual reopening | 3-18 | ADI-R, VABS-II/3 | y | DAS-II; WAIS-IV; WISC-IV; WAIS-IV; MSEL; Ravens-2; WPPSI-IV |

|  | **Contributing Sample Label** | **Contact Source** | **AFAR survey version/ administration mode** | **Catchment area** | **Data collection interval (d/m/2020)**^a^ | **COVID 19 response at the time of collection** | **Target age range (years)** | **Parent Interviews** | **ADOS** | **Child Cognitive Testing** |
| --- | --- | --- | --- | --- | --- | --- | --- | --- | --- | --- |
| **USA, Missouri** | TC | C | E 0.4.0; ReDCap | Missouri, Kansas Illinois | 06/11-10/20^a^ | No state-directed response specific to COVID-19; decisions about lockdowns and other restrictions/mandates varied considerably by city and county | 3-17 | ABAS | y | DAS-II; KBIT-2; WISC-IV/V; WAIS; WPPSI-IV |
| **Canada, Ontario** | POND-CMH | R | E 0.4.0; RedCap | South Ontario (Toronto, Hamilton, London, Kingston areas) | 5/14-8/27^a^ | 3/17-8/15 Lock down order for 100% non-essential workers | 3-21 | ADI-R, ABAS, K-SADS, PICS | y | WASI or other Wechsler tests, Standford Binet, Mullen |
| **Greece** | UAth | C | G 0.4.0, Google Forms; phone interviews | Athens metropolitan area (~75%) and west/central Greece + Islands (25%). | 05/06-06/25 | 03/11-5/15: Lockdown order for 100% of non-essential workforce followed by a gradual re-opening of schools and special services | 3-18 | ADI-R | y | Ravens; WISC-III |
| **Ireland** | TCD | C, R, National Advocacy^d^ | E 0.4.0^e^; Qualtrics | Dublin Metropolitan area, East Ireland | 06/09-08/17 | 03/20-05/20: Lockdown for 100% of non-essential workforce  06/08 -08/31: Gradual reopening | 3-20 | ADI-R^c^, VABS-II^c^, semi-structured psychiatric interviews | subset only | WISC |

|  | **Contributing Sample Label** | **Contact Source** | **AFAR survey version/ administration mode** | **Catchment area** | **Data collection interval (d/m/2020)**^a^ | **COVID 19 response at the time of collection** | **Target age range (years)** | **Parent Interviews** | **ADOS** | **Child Cognitive Testing** |
| --- | --- | --- | --- | --- | --- | --- | --- | --- | --- | --- |
| **Italy** | UBA | C | I 0.5.1; ReDCap | Puglia, South of Italy | 07/20-09/15 | 03/09-05/04: Lockdown order for 100% non-essential, followed by nearly full reopening (mask obligatory) ~ September | 3-18 | ADI-R, VABS-II/3 | y | WISC-IV; WPPSI-III; WAIS; Leiter-R/III; Merrill-Palmer; PEP-III |
|  | UCA | C |  | South Sardinia, Cagliari Oristano provinces | 07/10-09/10 |  | 3-18 | ADI-R, VABS-II/3; KSADS-PL | y | Griffiths-III; WAIS-IV; WISC-III/IV; WPPSI-III; Leiter-R |
|  | UONPI-LO | C |  | Lodi and surroundings, Lombardy | 07/02-09/17 |  | 3-18 | ADI-R, VABS-I | y | Leiter-R; WPPSI-III; WISC-IV |
|  | UFII | C |  | Napoli metropolitan area, Campania | 07/22-09/23 |  | 3-18 | ADI-R, VABS-II/3, KSADS-PL | y | Griffiths; Leiter; WISC-IV |
|  | UTV | C, R |  | Rome, Center-South of Italy | 06/20-09/20 |  | 3-18 | ABAS-II; VABS- II | y | WPPSI-III; WISC-IV; Leiter; PEP-III |
|  | USS | C, R |  | North Sardinia, Sassari and Olbia provinces | 07/28-09/21 |  | 3-17 | ADI-R, VABS-II/3 or ABAS | y | WPPSI-III; WISC-IV; GMDS-R; Leiter-R, Ravens |
|  | SMF | C |  | Tuscany and Center of Italy | 07/25-08/22 |  | 3-18 | ADI-R, VABS-II/3, KSADS-PL | y | WPPSI-III; WISC-IV; GMDS-R; Griffiths-III |

^a^Indicates actual data collection interval; for 4 samples (TC, UCSF, CMI-AC, POND-CMH) only data from children's survey completed within 2 weeks from the data collection interval of 90% of that sample were included in analyses. See Figure 1 for the data collection intervals of the selected datasets by sample. ^b^The latest 0.5.1 version was used as soon as it was available, a subset of children was administered an earlier version. ^c^ Only administered for a subset of the sample. ^d^AsIAm (https://asiam.ie/). ^e^Culturally adapted to the Irish population. Abbreviations: CMI-AC: Child Mind Institute-Autism Center, New York City; CMI-HBN:CMI-Healthy Brain Network, New York City; TC:Thompson Center, Columbia; UCSF:University of California San Francisco, San Francisco; CADB:Center for Autism and Developing Brain, Weill Cornell Medical College/New York Presbyterian Hospital, White Plains; UAth:University of Athens, National & Kapodistrian University of Athens, School of Medicine, First Department of Pediatrics, Unit of Developmental and Behavioral Pediatrics. “Aghia Sophia” Children’s Hospital, Athens, Greece; POND-CMH:Province of Ontario Neurodevelopmental Network, COVID Mental Health collaboration, Ontario; TCD:Trinity College Dublin, Dublin; UCA: University of Cagliari, Child & Adolescent Neuropsychiatry Unit, A.Cao Paediatric Hospital, G.Brotzu, Cagliari, Italy; UBA:University Bari, Child Neuropsychiatry Unit, Policlinic of Bari, Italy; UFII: University of Naples Federico II, Child and Adolescent Neuropsychiatry Unit, Naples, Italy; SMF:IRCCS Stella Maris Foundation, Pisa (Calambrone), Italy; UTV: UniversityTor Vergata, Rome, Italy; USS: University of Sassari, Child Neuropsychiatry Unit, Azienda Ospedaliero-Universitaria, Sassari; UONPI-LO:Unita' Operativa di Neuropsichiatria dell’ Infanzia e dell' adolescenza, Lodi, Italy; C: Clinic Contact, R: Active or Prior Research Contacts; E: English, G: Greek, I: Italian; Y: yes; ADI-R: Autism Diagnostic Interview- Revised (Rutter, Le Couteur, & Lord, 2003); KSADS: (Kaufman et al., 1997); VABS-II: Vineland-II (Sparrow et al., 2008); VABS-3: Vineland-3 (Sparrow, Cicchetti, & Saulnier, 2016), Adaptive Behavior Assessment Scale, Third Edition (ABAS-III; Harrison & Oakland, 2015); DAS-II: Differential Ability Scales, Second Edition (Elliot, 2007); MSEL:Mullen Scales of Early Learning (Mullen, 1995); WASI-2:Wechsler Abbreviated Scale of Intelligence, Second Edition (Wechsler, 2011); BAYLEY III; Ravens:Ravens Standard Progressive Matrices and Colored Progressive Matrices (Raven, Raven, & Court, J. H., 1998b); WISC-III:Wechsler Intelligence Scale for Children, Third Edition (Wechsler, 1991); WISC-IV:Wechsler Intelligence Scale for Children, fourth edition, Italian version, (Wechsler, 2004); WPPSI-III: Wechsler Preschool and Primary Scale of Intelligence-third edition; Leiter: Leiter International Performance Scale Revised-Visualization and Reasoning battery (Roid et al., 1997); Merrill-Palmer:Merrill-Palmer–Revised Scales of Development (Roid, 2004); PEP-III: Psychoeducational Profile-third edition (Schopler et al., 2005); GMDS-R:Griffiths Mental Development Scales (Griffiths, 1996); GMDS-ER:Griffiths Mental Development Scale (Luiz D et al. 2004).

**eTable 2: Clinical Characteristics of the aggregate dataset and 15 contributing samples.**

|  | **Aggregate** | **CMI-AC** | **CMI-HBN** | **CADB** | **TC** | **UCSF** | **POND-CMH** | **TCD** | **UAth** | **UBA** | **UCA** | **UONPI-LO** | **UFII** | **SMF** | **UTV** | **USS** |
| --- | --- | --- | --- | --- | --- | --- | --- | --- | --- | --- | --- | --- | --- | --- | --- | --- |
|  | N=1,275 | n=117 | n=87 | n=72 | n=44 | n=30 | n=300 | n =169 | n=44 | n=64 | n=88 | n=50 | n=37 | n=82 | n=53 | n=38 |
| **Neuropsychiatric Diagnoses, # (%)** |  |  |  |  |  |  |  |  |  |  |  |  |  |  |  |  |
| ASD | 1,004 (79) | 51 (44) | 87 (100) | 72 (100) | 29 (66) | 21 (70) | 146 (49) | 151 (89) | 44 (100) | 64 (100) | 82 (93) | 50 (100) | 37 (100) | 82 (100) | 53 (100) | 35 (92) |
| ADHD | 484 (38) | 81 (69) | 75 (86) | 4 (6) | 24 (55) | 9 (30) | 129 (43) | 37 (22) | 13 (30) | 20 (31) | 43 (49) | 0 | 3 (8) | 31 (38) | 4 (8) | 11 (29) |
| Intellectual Disability | 132 (10) | 0 | 1 (1) | 3 (4) | 1 (2) | 0 | 11 (4) | 15 (9) | 5 (11) | 7 (11) | 49 (56) | 0 | 9 (24) | 1 (1) | 6 (11) | 24 (63) |
| Borderline Intellectual Functioning | 2 (0) | 0 | 0 | 0 | 1 (2) | 0 | 0 | 0 | 0 | 0 | 0 | 0 | 0 | 0 | 1 (2) | 0 |
| Global Developmental Delay | 4 (0) | 0 | 0 | 0 | 0 | 0 | 0 | 0 | 0 | 0 | 4 (5) | 0 | 0 | 0 | 0 | 0 |
| Anxiety Disorder | 111 (9) | 30 (26) | 37 (43) | 3 (4) | 6 (14) | 6 (20) | 4 (1) | 6 (4) | 0 | 3 (5) | 5 (6) | 0 | 0 | 10 (12) | 1 (2) | 0 |
| Language, Learning, or Other NDD | 94 (7) | 6 (5) | 22 (25) | 3 (4) | 11 (25) | 4 (13) | 3 (1) | 2 (1) | 2 (5) | 13 (20) | 15 (17) | 0 | 0 | 11 (13) | 2 (4) | 0 |
| Disruptive, Impulse-Control, or Conduct Disorders | 73 (6) | 14 (12) | 12 (14) | 1 (1) | 7 (16) | 0 | 0 | 5 (3) | 1 (2) | 0 | 16 (18) | 0 | 0 | 13 (16) | 1 (2) | 3 (8) |
| Obsessive Compulsive Disorders | 30 (2) | 1 (1) | 5 (6) | 0 | 0 | 0 | 15 (5) | 1 (1) | 1 (2) | 0 | 2 (2) | 0 | 0 | 5 (6) | 0 | 0 |
| Mood Disorders | 23 (2) | 5 (4) | 5 (6) | 0 | 4 (9) | 1 (3) | 0 | 0 | 0 | 1 (2) | 7 (8) | 0 | 0 | 0 | 0 | 0 |
| Motor or Tic Disorders | 19 (1) | 3 (3) | 5 (6) | 0 | 1 (2) | 0 | 0 | 2 (1) | 0 | 3 (5) | 4 (5) | 0 | 0 | 1 (1) | 0 | 0 |
| Trauma, Psychosis, Substance Use & Personality Disorders | 16 (1) | 1 (1) | 6 (7) | 0 | 2 (5) | 0 | 0 | 1 (1) | 0 | 0 | 6 (7) | 0 | 0 | 0 | 0 | 0 |
|  | **Aggregate** | **CMI-AC** | **CMI-HBN** | **CADB** | **TC** | **UCSF** | **POND-CMH** | **TCD** | **UAth** | **UBA** | **UCA** | **UONPI-LO** | **UFII** | **SMF** | **UTV** | **USS** |
| **Neuropsychiatric Diagnoses, # (%)** |  |  |  |  |  |  |  |  |  |  |  |  |  |  |  |  |
| Known Genetic/Medical Conditions | 26 (2) | 0 | 0 | 0 | 0 | 0 | 9 (2) | 3 (2) | 7 (16) | 5 (8) | 2 (2) | 0 | 0 | 0 | 0 | 0 |
| **ASD w or w/o Comorbidities, N (%)** |  |  |  |  |  |  |  |  |  |  |  |  |  |  |  |  |
| ASD w/o Comorbidities | 570 (57) | 16 (31) | 4 (5) | 59 (82) | 6 (21) | 15 (71) | 134 (92) | 122 (81) | 20 (45) | 30 (47) | 10 (12) | 50 (100) | 25 (68) | 27 (33) | 40 (75) | 12 (34) |
| ASD + 1 Comorbidity | 232 (23) | 23 (45) | 22 (25) | 12 (17) | 10 (34) | 4 (19) | 12 (8) | 15 (10) | 19 (43) | 16 (25) | 27 (33) | 0 (0) | 12 (32) | 36 (44) | 12 (23) | 12 (34) |
| ASD > 1 Comorbidities | 202 (20) | 12 (24) | 61 (70) | 1 (1) | 13 (45) | 2 (10) | 0 (0) | 14 (9) | 5 (11) | 18 (28) | 45 (55) | 0 (0) | 0 (0) | 19 (23) | 1 (2) | 11 (31) |
| **Full-scale IQ ^b^** |  |  |  |  |  |  |  |  |  |  |  |  |  |  |  |  |
| Mean (SD) | 92.9 (23.5) | 104.8 (17.3) | 98.9 (17.5) | 85.8 (28.6) | 88.5 (13.9) | 108.8 (15.3) | 92 (23.5) | - | 105 (20.4) | 92.9 (26.6) | 83.7 (22.2) | 83.4 (17.9) | 77.2 (20.2) | 94.3 (21.7) | 77.1 (29.6) | 58.7 (33.3) |
| Range | 2-158 | 67-158 | 52-139 | 15-143 | 52-132 | 84-140 | 2-142 | - | 70-121 | 44-139 | 40-124 | 43-123 | 50-115 | 48-137 | 25-128 | 16-110 |
| *N* | 891^b^ | 117 | 87 | 72 | 44 | 29 | 300 | - | 5 | 33 | 41 | 34 | 12 | 82 | 18 | 15 |
| **Verbal IQ** |  |  |  |  |  |  |  |  |  |  |  |  |  |  |  |  |
| Mean (SD) | 95.6 (24.9) | 107.4 (17.4) | 103.4 (17.4) | 82.8 (33.2) | 91.6 (13.5) | 110.2 (20.1) | 91.7 (26.3) | - | 97.8 (17.5) | 101.5 (24.8) | 76 (20) | - | 82.4 (16.8) | 93.7 (24.5) | - | 82.6 (25.6) |
| Range | 3-170 | 70-170 | 62-139 | 9-136 | 59-121 | 77-155 | 3-160 | - | 70-117 | 54-138 | 26-108 | - | 62-110 | 50-148 | - | 45-124 |
| *N* | 717 | 116 | 87 | 69 | 39 | 25 | 234 | - | 5 | 29 | 11 | - | 9 | 81 | - | 11 |

|  | **Aggregate** | **CMI-AC** | **CMI-HBN** | **CADB** | **TC** | **UCSF** | **POND-CMH** | **TCD** | **UAth** | **UBA** | **UCA** | **UONPI-LO** | **UFII** | **SMF** | **UTV** | **USS** |
| --- | --- | --- | --- | --- | --- | --- | --- | --- | --- | --- | --- | --- | --- | --- | --- | --- |
| **Non-verbal IQ** |  |  |  |  |  |  |  |  |  |  |  |  |  |  |  |  |
| Mean (SD) | 94.2 (24.8) | 104.4 (18.6) | 101.7 (16.7) | 89.9 (27.4) | 92 (14.7) | 106.9 (14.4) | 93.2 (26.2) | - | - | 89.2 (27.9) | 71.9 (24.3) | - | 76 (25.4) | 101.7 (21.9) | 79.6 (27.4) | 80.6 (26.3) |
| Range | 2-162 | 58-162 | 52-140 | 16-158 | 60-139 | 88-140 | 2-147 | - | - | 19-148 | 27-115 | - | 36-119 | 50-148 | 36-153 | 38-120 |
| *N* | 833 | 117 | 87 | 72 | 44 | 25 | 237 | - | - | 57 | 42 | - | 19 | 82 | 35 | 11 |
| **ADOS-2** |  |  |  |  |  |  |  |  |  |  |  |  |  |  |  |  |
| Total CSS | 6.4 (2.4) | 5.2 (2.8) | 7.2 (2.8) | 7.7 (1.9) | 6.2 (2.7) | - | 7 (2.3) | - | - | - | - | - | - | 5.6 (1.3) | 6.8 (1.6) | - |
| N | 479 | 116 | 12 | 70 | 43 | - | 108 | - | - | - | - | - | - | 81 | 48 | - |
| RRB CSS | 6.2 (3.1) | 5.1 (3.3) | 6.4 (2.6) | 8 (1.7) | - | - | - | - | - | - | - | - | - | - | - | - |
| N | 198 | 116 | 12 | 70 | - | - | - | - | - | - | - | - | - | - | - | - |
| Social Affect CSS | 6.3 (2.6) | 5.6 (2.6) | 7.4 (2.7) | 7.4 (2.1) | - | - | - | - | - | - | - | - | - | - | - | - |
| N | 198 | 116 | 12 | 70 | - | - | - | - | - | - | - | - | - | - | - | - |
| Expressive Language score | 6.2 (2.2) | 7.8 (0.5) | 7.8 (0.4) | 6 (2.2) | 7.7 (0.6) | 7.3 (1.2) | 6.1 (2.2) | - | - | 5.5 (2.5) | 5.6 (2.4) | 4.8 (1.9) | 3.9 (2.4) | 6.6 (1.9) | 4.7 (2.5) | 5.4 (2.1) |
| N | 730 | 116 | 12 | 72 | 44 | 23 | 119 | - | - | 64 | 52 | 48 | 37 | 81 | 44 | 18 |
| **Adaptive Functioning, M(SD)** |  |  |  |  |  |  |  |  |  |  |  |  |  |  |  |  |
| Overall Adaptive Composite scores (VABS+ABAS) ^c^ | 71.3 (18.9) | 78.9 (11.8) | - | 73.2 (14.5) | 74.4 (17.1) | 77.9 (10.5) | 75.9 (16.8) | - | - | 48.8 (18.5) | - | - | 66.5 (30.3) | 69.3 (15.8) | 59 (16) | - |
| N | 674 | 100 | - | 64 | 41 | 26 | 238 | - | - | 64 | - | - | 33 | 61 | 47 | - |
|  | **Aggregate** | **CMI-AC** | **CMI-HBN** | **CADB** | **TC** | **UCSF** | **POND-CMH** | **TCD** | **UAth** | **UBA** | **UCA** | **UONPI-LO** | **UFII** | **SMF** | **UTV** | **USS** |
| **Child Behavior Checklist T scores, M (SD)** |  |  |  |  |  |  |  |  |  |  |  |  |  |  |  |  |
| Total problems | 64.1 (11.2) | 62.9 (9.1) | 61.8 (8.4) | 60.1 (10.9) | 68.5 (13) | 66.2 (9.4) | 66.2 (11.3) | - | - | 67 (7.1) | - | - | 59.1 (8.9) | 62.7 (10.6) | 60.6 (9.7) | 72.9 (7.3) |
| N | 748 | 105 | 80 | 66 | 40 | 19 | 247 | - | - | 13 | - | - | 37 | 82 | 42 | 14 |
| Externalizing problems | 59 (10.9) | 58.7 (9.9) | 56.6 (10.9) | 56.9 (11.9) | 64.3 (10.3) | 61.4 (9.6) | 60.6 (10.6) | - | - | 61 (6.4) | - | - | 55.4 (10.5) | 57.4 (11.3) | 55.6 (9.4) | 70.4 (7.2) |
| N | 716 | 105 | 80 | 66 | 40 | 19 | 215 | - | - | 13 | - | - | 37 | 82 | 42 | 14 |
| Internalizing problems | 62.3 (11.6) | 60 (11.4) | 59.8 (9.4) | 57.9 (11.2) | 67.5 (12.5) | 65.5 (10.4) | 64 (12.7) | - | - | 65.2 (7.6) | - | - | 58.5 (9.9) | 64.6 (10.4) | 60.4 (8.8) | 69.1 (9.9) |
| N | 749 | 106 | 80 | 66 | 40 | 19 | 247 | - | - | 13 | - | - | 37 | 82 | 42 | 14 |
| **Current Medication use, N (%)** |  |  |  |  |  |  |  |  |  |  |  |  |  |  |  |  |
| Yes | 489 (38) | 44 (38) | 32 (37) | 24 (33) | 27 (61) | 8 (27) | 166 (55) | 65 (38) | 2 (5) | 14 (22) | 50 (57) | 4 (8) | 5 (14) | 34 (41) | 4 (8) | 10 (26) |
| *N* | 1274 | 117 | 86 | 72 | 44 | 30 | 300 | 169 | 44 | 64 | 88 | 50 | 37 | 82 | 53 | 38 |
| **Time of collection difference (years), M (SD)** |  |  |  |  |  |  |  |  |  |  |  |  |  |  |  |  |
| IQ test administration | 2.2 (3.3) | 1.4 (1.2) | 1.5 (1.1) | 3.3 (1.9) | 1.1 (0.9) | 2.5 (1.6) | 3.3 (2.1) | - | 4.4 (1.2) | 1.4 (2.5) | 2.6 (2.7) | 2.5 (1.7) | 1.8 (1.8) | 1.7 (1.7) | 1.5 (1.4) | 2 (1.6) |
| ADOS-2 administration | 2.1 (2.2) | 1.6 (2.8) | 0.7 (0.5) | 2.5 (1.7) | 1.1 (0.9) | 3.3 (2.1) | 2.8 (2.1) | - | - | 1 (1.9) | 2.3 (3.1) | 2.5 (1.7) | 2.4 (2.2) | 1.8 (1.8) | 1.7 (1.1) | 4 (2) |
| VABS/ABAS administration | 2.2 (1.8) | 1.5 (1.1) | - | 2.5 (1.8) | 1.1 (0.5) | 2.3 (1.5) | 3.1 (2.2) | - | - | 1.5 (1.2) | - | - | 1.8 (1.6) | 2 (1.9) | 1.2 (1) | - |
| CBCL administration | 2 (2) | 1.4 (1.2) | 1.4 (1.2) | 2.4 (1.6) | 1.1 (0.7) | 2.7 (2) | 3.2 (2.3) | - | - | 1.5 (3) | - | - | 0.1 (0.7) | 1.8 (1.8) | 1.7 (1.8) | 1.1 (1.1) |

****** The Mean and Standard Deviation for the above clinical measures collected prior to the pandemic is shown for each contributing sample when the data was available for 5 or more participants. ^a^This includes any diagnoses and double counts those individuals who have more than one diagnoses. ^b^ Out of these n=891 total participants from the aggregate dataset with available FIQ quantitative scores, there are n= 759 with FIQ quantitative scores reported by each contributing samples (when available), n=15 were averaged from VIQ and NVIQ scores , n=63 were DQ scores and n=54 were imputed based on age and diagnosis in those samples with less than 15% data missing (see supplementary methods for more details on the imputation method used). ^c^ CMI-HBN, UCA and UONPI-LO administered the VABS to some children of their sample (n=5, n=19, n=50, respectively), however the overall composite scores were not available. Abbreviations: Autism Spectrum Disorder (ASD), Attention-Deficit/Hyperactivity Disorder (ADHD), IC/CD: Impulse-Control and Conduct Disorders; NDD: Neurodevelopmental disorders. Other abbreviations: see eTable1 for site name abbreviated labels.

**eTable 3: Participant characteristics of the aggregate dataset and 15 contributing samples.**

|  | **Aggregate** | **CMI-AC** | **CMI-HBN** | **CADB** | **TC** | **UCSF** | **POND-CMH** | **TCD** | **UAth** | **UBA** | **UCA** | **UONPI-LO** | **UFII** | **SMF** | **UTV** | **USS** |
| --- | --- | --- | --- | --- | --- | --- | --- | --- | --- | --- | --- | --- | --- | --- | --- | --- |
|  | N=1,275 | n=117 | n=87 | n=72 | n=44 | n=30 | n=300 | n =169 | n=44 | n=64 | n=88 | n=50 | n=37 | n=82 | n=53 | n=38 |
| **Child Demographics** |  |  |  |  |  |  |  |  |  |  |  |  |  |  |  |  |
| **Age, Years** |  |  |  |  |  |  |  |  |  |  |  |  |  |  |  |  |
| Mean (SD) | 11 (3.6) | 10.2 (2.2) | 12.3 (3.8) | 9.2 (3.3) | 10.1 (3.3) | 10.8 (3.8) | 12.1 (3.5) | 12 (3.3) | 10 (3.1) | 10.4 (3.9) | 11.6 (3.7) | 9.9 (3.1) | 8.7 (3.4) | 10.6 (3.5) | 10.2 (3.7) | 9.2 (3.5) |
| min-max | 5-21 | 6-15 | 6-21 | 5-19 | 6-17 | 5-18 | 5-18 | 5-19 | 5-16 | 5-18 | 5-18 | 5-16 | 5-17 | 5-19 | 5-17 | 5-16 |
| N | 1275 | 117 | 87 | 72 | 44 | 30 | 300 | 169 | 44 | 64 | 88 | 50 | 37 | 82 | 53 | 38 |
| **Sex^a^ #, (%)** |  |  |  |  |  |  |  |  |  |  |  |  |  |  |  |  |
| Males | 996 (78) | 89 (76) | 73 (84) | 60 (83) | 32 (73) | 19 (63) | 220 (73) | 136 (80) | 35 (80) | 52 (81) | 75 (85) | 43 (86) | 27 (73) | 66 (80) | 44 (83) | 25 (66) |
| Females | 277 (22) | 28 (24) | 14 (16) | 12 (17) | 12 (27) | 11 (37) | 79 (26) | 32 (19) | 9 (20) | 12 (19) | 13 (15) | 7 (14) | 10 (27) | 16 (20) | 9 (17) | 13 (34) |
| N | 1275 | 117 | 87 | 72 | 44 | 30 | 300 | 169 | 44 | 64 | 88 | 50 | 37 | 82 | 53 | 38 |
| **Gender Identity** |  |  |  |  |  |  |  |  |  |  |  |  |  |  |  |  |
| Boy/Man | 932 (77) | 89 (76) | 70 (81) | 60 (83) | - | 19 (63) | 219 (73) | 137 (81) | 35 (80) | 47 (80) | 69 (83) | 40 (82) | 25 (71) | 62 (78) | 41 (84) | 19 (59) |
| Girl/Woman | 251 (21) | 26 (22) | 14 (16) | 12 (17) | - | 11 (37) | 78 (26) | 30 (18) | 9 (20) | 10 (17) | 12 (14) | 6 (12) | 10 (29) | 16 (20) | 7 (14) | 10 (31) |
| Transgender | 3 (0) | 1 (1) | 0 | 0 | - | 0 | 1 (0) | 1 (1) | 0 | 0 | 0 | 0 | 0 | 0 | 0 | 0 |
| Non-binary | 4 (0) | 1 (1) | 2 (2) | 0 | - | 0 | 1 (0) | 0 | 0 | 0 | 0 | 0 | 0 | 0 | 0 | 0 |
| Missing | 14 (1) | 0 | 0 | 0 | - | 0 | 1 (0) | 1 (1) | 0 | 2 (3) | 2 (2) | 3 (6) | 0 | 1 (1) | 1 (2) | 3 (9) |
| N | 1204 | 117 | 86 | 72 | - | 30 | 300 | 169 | 44 | 59 | 83 | 49 | 35 | 79 | 49 | 32 |
|  | **Aggregate** | **CMI-AC** | **CMI-HBN** | **CADB** | **TC** | **UCSF** | **POND-CMH** | **TCD** | **UAth** | **UBA** | **UCA** | **UONPI-LO** | **UFII** | **SMF** | **UTV** | **USS** |
| **Hispanic^c^ Ethnicity** | 97 (15) | 35 (30) | 30 (34) | 19 (26) | 2 (5) | 1 (3) | - | - | - | 2 (4) | 2 (3) | 2 (5) | 0 | 1 (2) | 1 (3) | 2 (7) |
| N | 646 | 117 | 87 | 72 | 44 | 30 | - | - | - | 46 | 61 | 41 | 32 | 53 | 34 | 29 |
| **Educational Setting Prior to COVID-19^d^, #, (%)** |  |  |  |  |  |  |  |  |  |  |  |  |  |  |  |  |
| General education school w support | 636 (50) | 64 (55) | 31 (36) | 21 (29) | 25 (57) | 20 (67) | 130 (44) | 73 (43) | 22 (50) | 38 (59) | 46 (52) | 34 (68) | 18 (49) | 58 (71) | 33 (62) | 23 (61) |
| General Education classroom w/o support | 183 (14) | 9 (8) | 4 (5) | 1 (1) | 7 (16) | 2 (7) | 87 (29) | 21 (12) | 13 (30) | 8 (12) | 10 (11) | 3 (6) | 4 (11) | 10 (12) | 3 (6) | 1 (3) |
| Special Education classroom | 379 (30) | 43 (37) | 50 (57) | 48 (67) | 10 (23) | 6 (20) | 68 (23) | 43 (25) | 9 (20) | 13 (20) | 30 (34) | 12 (24) | 13 (35) | 11 (13) | 14 (26) | 9 (24) |
| Specialized program | 72 (6) | 1 (1) | 2 (2) | 2 (3) | 2 (5) | 2 (7) | 10 (3) | 32 (19) | 0 | 5 (8) | 2 (2) | 1 (2) | 2 (5) | 3 (4) | 3 (6) | 5 (13) |
| N | 1270 | 117 | 87 | 72 | 44 | 30 | 295 | 169 | 44 | 64 | 88 | 50 | 37 | 82 | 53 | 38 |
| **Caregiver/Household Demographics** |  |  |  |  |  |  |  |  |  |  |  |  |  |  |  |  |
| **Respondant Age, Years^e^** |  |  |  |  |  |  |  |  |  |  |  |  |  |  |  |  |
| Mean (SD) | 44.3 (6.5) | 43.5 (6.6) | 46.3 (7.3) | 42.5 (6.5) | - | - | - | 45.1 (6.2) | 43.3 (5.4) | 42.5 (6.4) | 45.8 (6.2) | 43.4 (6.1) | 40.6 (6.4) | 45.2 (6.1) | 44.6 (6.7) | 44.1 (5.8) |
| min-max | 24-70 | 26-61 | 32-70 | 30-68 | - | - | - | 31-59 | 30-56 | 26-55 | 32-60 | 31-61 | 24-59 | 30-62 | 29-56 | 31-58 |
| N | 1275 | 117 | 87 | 72 | - | - | - | 169 | 44 | 64 | 88 | 50 | 37 | 82 | 53 | 38 |
|  | **Aggregate** | **CMI-AC** | **CMI-HBN** | **CADB** | **TC** | **UCSF** | **POND-CMH** | **TCD** | **UAth** | **UBA** | **UCA** | **UONPI-LO** | **UFII** | **SMF** | **UTV** | **USS** |
| **Respondent Relationship to Child^f^, #, (%)** |  |  |  |  |  |  |  |  |  |  |  |  |  |  |  |  |
| Mother | 1,062 (86) | 100 (85) | 77 (89) | 65 (90) | - | 26 (87) | 278 (93) | 157 (93) | 37 (84) | 52 (81) | 69 (78) | 42 (84) | 32 (86) | 56 (68) | 41 (77) | 30 (79) |
| Father | 155 (13) | 17 (15) | 10 (11) | 7 (10) | - | 4 (13) | 18 (6) | 11 (7) | 7 (16) | 10 (16) | 17 (19) | 7 (14) | 5 (14) | 24 (29) | 11 (21) | 7 (18) |
| Other | 14 (1) | 0 | 0 | 0 | - | 0 | 4 (1) | 1 (1) | 0 | 2 (3) | 2 (2) | 1 (2) | 0 | 2 (2) | 1 (2) | 1 (3) |
| *N* | 1231 | 117 | 87 | 72 | - | 30 | 300 | 169 | 44 | 64 | 88 | 50 | 37 | 82 | 53 | 38 |
| **Urbanicity, #, (%)** |  |  |  |  |  |  |  |  |  |  |  |  |  |  |  |  |
| Large city | 392 (31) | 86 (74) | 46 (54) | 21 (29) | 3 (7) | 13 (43) | 121 (40) | 19 (11) | 24 (65) | 10 (16) | 12 (14) | 2 (4) | 3 (8) | 7 (9) | 21 (40) | 4 (11) |
| Town, Village, or Rural Area | 356 (28) | 3 (3) | 10 (12) | 21 (29) | 22 (50) | 4 (13) | 36 (12) | 98 (58) | 3 (8) | 24 (38) | 31 (36) | 31 (62) | 21 (57) | 29 (35) | 9 (17) | 14 (37) |
| Suburbs of a large city | 255 (20) | 21 (18) | 28 (33) | 26 (36) | 6 (14) | 9 (30) | 81 (27) | 52 (31) | 6 (16) | 9 (14) | 4 (5) | 1 (2) | 1 (3) | 4 (5) | 5 (9) | 2 (5) |
| Small city | 261 (21) | 7 (6) | 1 (1) | 4 (6) | 13 (30) | 4 (13) | 61 (20) | 0 | 4 (11) | 21 (33) | 40 (46) | 16 (32) | 12 (32) | 42 (51) | 18 (34) | 18 (47) |
| *N* | 1264 | 117 | 85 | 72 | 44 | 30 | 299 | 169 | 37 | 64 | 87 | 50 | 37 | 82 | 53 | 38 |
| **Essential Worker in the household, #, (%)** |  |  |  |  |  |  |  |  |  |  |  |  |  |  |  |  |
| Yes | 397 (31) | 30 (26) | 25 (29) | 20 (28) | 27 (61) | 17 (57) | 96 (32) | 72 (43) | 20 (45) | 7 (11) | 20 (23) | 10 (21) | 8 (22) | 20 (25) | 17 (32) | 8 (21) |
| *N* | 1271 | 117 | 87 | 72 | 44 | 30 | 300 | 169 | 44 | 63 | 88 | 48 | 37 | 81 | 53 | 38 |
|  | **Aggregate** | **CMI-AC** | **CMI-HBN** | **CADB** | **TC** | **UCSF** | **POND-CMH** | **TCD** | **UAth** | **UBA** | **UCA** | **UONPI-LO** | **UFII** | **SMF** | **UTV** | **USS** |
| **Household Composition, #, (%)** |  |  |  |  |  |  |  |  |  |  |  |  |  |  |  |  |
| Two Parents | 884 (69) | 80 (68) | 57 (66) | 51 (71) | 26 (59) | 25 (83) | 196 (65) | 111 (66) | 30 (68) | 52 (81) | 67 (76) | 44 (88) | 25 (68) | 56 (68) | 42 (79) | 22 (58) |
| One Parent | 187 (15) | 19 (16) | 7 (8) | 5 (7) | 6 (14) | 3 (10) | 57 (19) | 30 (18) | 11 (25) | 5 (8) | 10 (11) | 4 (8) | 4 (11) | 12 (15) | 7 (13) | 7 (18) |
| Multigeneration/Other Family members | 125 (10) | 11 (9) | 17 (20) | 7 (10) | 6 (14) | 2 (7) | 36 (12) | 12 (7) | 1 (2) | 4 (6) | 7 (8) | 1 (2) | 1 (3) | 12 (15) | 3 (6) | 5 (13) |
| Other | 79 (6) | 7 (6) | 6 (7) | 9 (12) | 6 (14) | 0 | 11 (4) | 16 (9) | 2 (5) | 3 (5) | 4 (5) | 1 (2) | 7 (19) | 2 (2) | 1 (2) | 4 (11) |
| *N* | 1275 | 117 | 87 | 72 | 44 | 30 | 300 | 169 | 44 | 64 | 88 | 50 | 37 | 82 | 53 | 38 |
| **Siblings, #, (%)** |  |  |  |  |  |  |  |  |  |  |  |  |  |  |  |  |
| Yes | 843 (79) | 74 (75) | 48 (75) | 41 (73) | 26 (81) | 25 (89) | 210 (83) | 116 (82) | 33 (80) | 47 (82) | 55 (71) | 36 (75) | 21 (72) | 49 (72) | 40 (82) | 22 (76) |
| *N* | 1071 | 99 | 64 | 56 | 32 | 28 | 253 | 141 | 41 | 57 | 77 | 48 | 29 | 68 | 49 | 29 |
| **Government assistance, #, (%)** |  |  |  |  |  |  |  |  |  |  |  |  |  |  |  |  |
| Yes | 312 (25) | 4 (3) | 4 (5) | 1 (1) | 5 (11) | 0 | 36 (12) | 23 (14) | 21 (49) | 28 (44) | 51 (58) | 28 (58) | 16 (43) | 29 (36) | 36 (68) | 30 (81) |
| *N* | 1268 | 116 | 87 | 72 | 44 | 30 | 300 | 169 | 43 | 63 | 88 | 48 | 37 | 81 | 53 | 37 |
|  | **Aggregate** | **CMI-AC** | **CMI-HBN** | **CADB** | **TC** | **UCSF** | **POND-CMH** | **TCD** | **UAth** | **UBA** | **UCA** | **UONPI-LO** | **UFII** | **SMF** | **UTV** | **USS** |
| **COVID-19 Health/Job Impact** |  |  |  |  |  |  |  |  |  |  |  |  |  |  |  |  |
| Child Two week COVID-19 Exposure, #, (%) |  |  |  |  |  |  |  |  |  |  |  |  |  |  |  |  |
| None | 1,223 (96) | 98 (84) | 73 (84) | 67 (93) | 39 (89) | 30 (100) | 297 (99) | 169 (100) | 44 (100) | 64 (100) | 87 (99) | 46 (92) | 37 (100) | 81 (99) | 53 (100) | 38 (100) |
| Exposure to person with diagnosis | 33 (3) | 9 (8) | 8 (9) | 5 (7) | 5 (11) | 0 | 3 (1) | 0 | 0 | 0 | 0 | 2 (4) | 0 | 1 (1) | 0 | 0 |
| Exposure to person with symptoms | 19 (1) | 10 (9) | 6 (7) | 0 | 0 | 0 | 0 | 0 | 0 | 0 | 1 (1) | 2 (4) | 0 | 0 | 0 | 0 |
| *N* | 1275 | 117 | 87 | 72 | 44 | 30 | 300 | 169 | 44 | 64 | 88 | 50 | 37 | 82 | 53 | 38 |
| **Family members COVID19 diagnosed, #, (%)** |  |  |  |  |  |  |  |  |  |  |  |  |  |  |  |  |
| No diagnosis | 1,213 (95) | 95 (81) | 74 (85) | 63 (88) | 43 (98) | 30 (100) | 297 (99) | 168 (99) | 44 (100) | 62 (97) | 87 (99) | 44 (88) | 37 (100) | 80 (98) | 52 (98) | 37 (97) |
| Yes, non-household member | 40 (3) | 15 (13) | 7 (8) | 4 (6) | 0 | 0 | 1 (0) | 1 (1) | 0 | 2 (3) | 1 (1) | 5 (10) | 0 | 2 (2) | 1 (2) | 1 (3) |
| Yes, household member | 22 (2) | 7 (6) | 6 (7) | 5 (7) | 1 (2) | 0 | 2 (1) | 0 | 0 | 0 | 0 | 1 (2) | 0 | 0 | 0 | 0 |
| *N* | 1275 | 117 | 87 | 72 | 44 | 30 | 300 | 169 | 44 | 64 | 88 | 50 | 37 | 82 | 53 | 38 |
|  | **Aggregate** | **CMI-AC** | **CMI-HBN** | **CADB** | **TC** | **UCSF** | **POND-CMH** | **TCD** | **UAth** | **UBA** | **UCA** | **UONPI-LO** | **UFII** | **SMF** | **UTV** | **USS** |
| **Family members Health/Job Impact^g^, #, (%)** |  |  |  |  |  |  |  |  |  |  |  |  |  |  |  |  |
| Lost job/Reduced income | 237 (19) | 23 (20) | 14 (16) | 12 (17) | 5 (11) | 6 (20) | 85 (28) | 17 (10) | 5 (11) | 10 (16) | 20 (23) | 7 (14) | 4 (11) | 9 (11) | 11 (21) | 9 (24) |
| Self-quarantine | 95 (7) | 10 (9) | 14 (16) | 13 (18) | 8 (18) | 0 | 33 (11) | 2 (1) | 1 (2) | 2 (3) | 4 (5) | 3 (6) | 0 | 2 (2) | 2 (4) | 1 (3) |
| Physical illness/Hospitalized | 45 (4) | 14 (12) | 10 (11) | 3 (4) | 0 | 0 | 10 (3) | 1 (1) | 0 | 1 (2) | 1 (1) | 3 (6) | 1 (3) | 1 (1) | 0 | 0 |
| Passed away | 6 (0) | 2 (2) | 1 (1) | 0 | 0 | 0 | 2 (1) | 0 | 0 | 0 | 0 | 1 (2) | 0 | 0 | 0 | 0 |
| None of the above | 892 (70) | 68 (58) | 48 (55) | 44 (61) | 31 (70) | 24 (80) | 170 (57) | 149 (88) | 38 (86) | 51 (80) | 63 (72) | 36 (72) | 32 (86) | 70 (85) | 40 (75) | 28 (74) |
| N | 1275 | 117 | 87 | 72 | 44 | 30 | 300 | 169 | 44 | 64 | 88 | 50 | 37 | 82 | 53 | 38 |

^a^Question on sex included three response options: male, female and other. n=1 respondent in TCD indicated "Other", n=1 from POND-CMH chose not to answer. ^b^ Gender identity information was not collected at TC (Thompson Center). Among the n=3 respondents selecting transgender: n=1 indicated Trans boy/man and n=2 Trans girl/woman. ^c^The question on Hispanic ethnicity was not included in the POND-CMH, TCD and UAth sample's surveys. ^d^ Special Education classroom refers to public or private schools; Specialized program refers to center- or home-based applied behavioral analysis programs or residential settings and alike. ^e^ Respondent age question was not included in the POND-CMH, TC and UCSF samples' survey. ^f^Respondent relationship to the child was not included in the TC site. ^g^ Self-quarantine summarizes answers indicating self-quarantine with and without symptoms. Other abbreviations: see eTable 1 for site name abbreviated labels.

**eTable 4: Ancestry of the aggregate dataset and 15 contributing samples.**

|  | **Aggregate** | **CMI-AC** | **CMI-HBN** | **CADB** | **TC ^a^** | **UCSF ^a^** | **POND-CMH ^a^** | **TCD ^a^** | **UAth** | **UBA** | **UCA** | **UONPI-LO** | **UFII** | **SMF** | **UTV** | **USS** |
| --- | --- | --- | --- | --- | --- | --- | --- | --- | --- | --- | --- | --- | --- | --- | --- | --- |
| N (%) | N=1,275 | n=117 | n=87 | n=72 | n=44 | n=30 | n=300 | n =169 | n=44 | n=64 | n=88 | n=50 | n=37 | n=82 | n=53 | n=38 |
| **European/British ^b^** | 716 (56) | 27 (23) | 24 (28) | 10 (14) | 0 | 0 | 151 (50) | 156 (92) | 41 (93) | 55 (86) | 56 (64) | 31 (62) | 31 (84) | 64 (78) | 47 (89) | 23 (61) |
| **Asian** | 45 (4) | 6 (5) | 2 (2) | 6 (8) | 0 | 4 (13) | 20 (7) | 2 (2) | 0 | 0 | 0 | 3 (6) | 0 | 0 | 1 (2) | 0 |
| **African** | 16 (1) | 4 (3) | 4 (5) | 4 (6) | 0 | 0 | 1 (0) | 2 (1) | 0 | 0 | 0 | 1 (2) | 0 | 0 | 0 | 0 |
| **Central / South American / Caribbean** | 14 (1) | 6 (5) | 3 (3) | 2 (3) | 0 | 0 | 3 (1) | 0 | 0 | 0 | 0 | 0 | 0 | 0 | 0 | 0 |
| **Middle Eastern** | 8 (1) | 4 (3) | 2 (2) | 1 (1) | 0 | 0 | 0 | 0 | 1 (2) | 0 | 0 | 0 | 0 | 0 | 0 | 0 |
| **Indigenous** | 11 (1) | 1 (1) | 0 | 1 (1) | 1 (2) | 0 | 5 (2) | 0 | 1 (2) | 0 | 1 (1) | 1 (2) | 0 | 0 | 0 | 0 |
| **Other ^c^** | 99 (8) | 4 (3) | 3 (3) | 1 (1) | 0 | 0 | 71 (24) | 0 | 1 (2) | 2 (3) | 7 (8) | 1 (2) | 2 (5) | 2 (2) | 1 (2) | 4 (11) |
| **Mixed** | 204 (16) | 61 (52) | 46 (53) | 40 (56) | 0 | 0 | 38 (13) | 0 | 0 | 1 (2) | 3 (3) | 3 (6) | 0 | 7 (9) | 3 (6) | 2 (5) |
| **Missing/Unknown** | 162 (13) | 4 (3) | 3 (3) | 7 (10) | 43 (98) | 26 (87) | 11 (4) | 9 (5) | 0 | 6 (9) | 21 (24) | 10 (20) | 4 (11) | 9 (11) | 1 (2) | 9 (24) |
| ^a^For the TC, UCSF, TCD and POND-CMH samples the response/options referring to ancestry were reworded slightly differently from AFAR. Whenever possible they were remapped into the AFAR structure. Specifically, for the TC and UCSF samples "American Indian," "Asian" and "African American" were mapped into the AFAR categories "Indigenous", "Asian" and "African"; the response "White" could not be obviously remapped into AFAR ancestry categories and was counted under "Missing/Unknown." For TCD "Irish" / "Irish Traveller," "African" and "Other Asian" were mapped into the "European/British," "African" and "Asian" ancestry categories of AFAR, respectively; the responses "Any other white background", "Any other black background" and "Chinese" could not be obviously mapped into AFAR ancestry categories and thus were listed under "Missing/Unknown." For the POND-CMH sample the responses "European Origins," "Caribbean origins" and "Latin, Central, South American origins," "African origins", "Asian origins," "North American Aboriginal origins" and "Oceania origins", "Other north American origins" and "Other" were mapped into "European/British," "Central/South American/Caribbean", "African," "Asian," "Indigenous," and "Other," respectively. ^b^ Includes European, British/Irish, Australian, New Zealand, North American (non-native). ^c^The respondent selected "Other" and was not further specified. Abbreviations: see eTable 1 for site name abbreviated labels. | | | | | | | | | | | | | | | | |

**eTable 5: Factor Loadings by domain examined.**

| **Domain/Item** | **Factor/Factor Loadings** | | | | |
| --- | --- | --- | --- | --- | --- |
|  | **F1** | **F2** | **F3** | **F4** | **F5** |
| Adaptive Living skills |  |  |  |  |  |
| Play/entertain self | **0.64** |  |  |  |  |
| Structure own activities | **0.77** |  |  |  |  |
| Self-care | **0.80** |  |  |  |  |
| Meal/food behaviors | **0.70** |  |  |  |  |
| Restricted and Repetitive Behaviors/Interests |  |  |  |  |  |
| Sensory seeking | **0.72** | -0.01 |  |  |  |
| Repetitive motor mannerisms | **0.80** | -0.04 |  |  |  |
| Rituals/routines | **0.50** | 0.36 |  |  |  |
| Makes family keep routines/rituals | -0.01 | **0.76** |  |  |  |
| Restricted interests | 0.20 | **0.29** |  |  |  |
| *Adjusts to change* | *0.11* | *0.02* |  |  |  |
| COVID-19 Worries |  |  |  |  |  |
| Worried about own infection | **0.92** | -0.07 |  |  |  |
| Worried about others’ infection | **0.77** | 0.07 |  |  |  |
| Worried about own physical health | **0.51** | 0.29 |  |  |  |
| *Worried about own mental health* | *0.01* | ***0.99*** |  |  |  |
| Reading/talking about/watching news about COVID-19 | **0.44** | 0.10 |  |  |  |
| *Positive change* | *0.00* | *0.01* | ***0.70*** |  |  |
| *Time outside* | *-0.06* | *0.02* | *-0.05* |  |  |
| Restriction stress | 0.05 | **0.60** | 0.25 |  |  |
| Cancellation difficulty | 0.01 | **0.90** | -0.06 |  |  |
| Financial difficulty | **0.37** | 0.17 | 0.06 |  |  |
| Living difficulty | **0.82** | 0.03 | -0.09 |  |  |
| Food security worry | **0.90** | -0.03 | 0.06 |  |  |
| *Hopefully End* | *0.05* | *-0.19* | *0.09* |  |  |
| Co-Occurring Problem Behaviors |  |  |  |  |  |
| Hyperactivity | 0.02 | -0.02 | **0.75** |  |  |
| Off task behaviors | 0.05 | 0.04 | **0.59** |  |  |
| Angry/Losing temper | **0.83** | 0.01 | 0.08 |  |  |
| Verbal aggression | **0.92** | -0.02 | -0.11 |  |  |
| Physical aggression | **0.62** | 0.02 | 0.16 |  |  |
| *Deliberately injuring self* | *0.00* | *0.19* | ***0.27*** |  |  |
| Disobedient/arguing | **0.63** | 0.09 | 0.10 |  |  |
| Crying easily | 0.18 | **0.37** | 0.14 |  |  |
| Social worries | 0.04 | **0.61** | -0.05 |  |  |
| Problems separating | -0.07 | **0.63** | 0.07 |  |  |
| Excessive fear | 0.02 | **0.77** | 0.03 |  |  |
| Weekday bedtime | **0.78** | -0.03 | -0.05 |  |  |
| Weekend bedtime | **0.91** | 0.00 | 0.04 | -0.03 | -0.02 |
| Weekday hours of sleep | -0.15 | **0.76** | -0.06 | -0.01 | -0.07 |
| Weekend hours of sleep | 0.06 | **0.95** | 0.03 | 0.00 | 0.02 |
| Difficulties falling asleep | 0.05 | 0.04 | -0.01 | -0.07 | **0.74** |
| Night waking | -0.11 | -0.12 | 0.00 | 0.09 | **0.64** |
| Exercise | 0.09 | -0.04 | 0.03 | **0.53** | 0.03 |
| Time outdoors | -0.01 | 0.01 | 0.02 | **0.84** | 0.00 |
| Time watching television | 0.10 | 0.03 | **0.27** | 0.08 | 0.11 |
| Time on social media | 0.07 | 0.04 | **0.55** | 0.02 | 0.11 |
| Time on video games | 0.10 | 0.09 | **0.33** | 0.11 | 0.08 |
| Online interactions with peers^a^ | 0.00 | -0.01 | **0.77** | 0.00 | -0.02 |
| Online interactions with adults^a^ | -0.11 | -0.04 | **0.54** | -0.01 | -0.10 |
| ^a^Item missed for three participants (n=2 UAth, n=1 POND-CMH)  Legend: The Grey and italicized text indicates items excluded in subsequent confirmatory factor analysis (CFA); items were excluded if their factor loadings resulting from EFA were below 0.03 (n=3 items) or if they resulted as a single item factor (n=2) or based on theoretical plausibility (n=1). See eTable 6 for CFA goodness-of-fit indices. Abbreviations: EFA, Exploratory Factor Analysis; F=Factor. Abbreviations: UAth: University of Athens, National & Kapodistrian University of Athens, School of Medicine, First Department of Pediatrics, Unit of Developmental and Behavioral Pediatrics. “Aghia Sophia” Children’s Hospital, Athens, Greece; POND-CMH:Province of Ontario Neurodevelopmental Network, COVID Mental Health collaboration. | | | | | |

**eTable 6: Goodness of Fit Indices for Exploratory and Confirmatory Factor Analyses.**

| **Domain** | **Analysis Type: Result** | **χ^2^** | **RMSEA** | **TLI** | **CFI** |
| --- | --- | --- | --- | --- | --- |
| **Adaptive Living Skills** | EFA: 1 factor | <0.01 | 0.03 | 0.97 |  |
|  | CFA-P: 1 factor | 0.20 | 0.03 | 1.00 | 1.00 |
|  | CFA-S: 1 factor | 0.13 | 0.03 | 1.00 | 1.00 |
| **RRB** | EFA: 2 factors, 6 items | <0.66 | 0.05 | 1.01 |  |
|  | CFA-P: 2 factors, 5 items | 0.03 | 0.05 | 1.00 | 1.00 |
|  | CFA-S: 2 factors, 5 items | <0.01 | 0.07 | 0.99 | 1.00 |
| **Co-Occurring Problem Behaviors** | EFA: 3 factors, 11 items | <0.00 | 0.06 | 0.95 |  |
|  | CFA-P: 3 factors, 10 items | <0.01 | 0.06 | 0.99 | 1.00 |
|  | CFA-S: 3 factors, 10 items | <0.01 | 0.07 | 0.99 | 0.99 |
| **Daily Behaviors** | EFA: 5 factors, 13 items | <0.00 | 0.09 | 0.83 |  |
|  | CFA-P: 5 factors, 13 items | <0.00 | 0.07 | 0.99 | 0.99 |
|  | CFA-S: 5 factors, 13 items | <0.01 | 0.11 | 0.99 | 1.00 |
| **COVID Worries^a^** | EFA: 2 factors | <0.35 | 0.00 | 1.00 |  |
|  | CFA-P: 1 factor, 4 items | 0.58 | 0.00 | 1.00 | 1.00 |
| **Life Changes^a^** | EFA: 3 factors | <0.97 | 0.03 | 0.98 |  |
|  | CFA-P: 2 factors, 5 items | 0.47 | 0.00 | 1.00 | 1.00 |

^a^These domains did not have Prior and Current time points; therefore, their EFA was made up of split-half 1 current scores (n=636) and CFA-P conducted on split-half 2 (n=637) current scores. Abbreviations: EFA, Primary EFA conducted on split-half sample 1 (n=636) using prior three month scores; CFA-P, Primary CFA conducted on split-half sample 2 (n=637) using the prior three months to COVID-19 pandemic scores; CFA-S, Secondary CFA was conducted on the whole sample using the current scores (n= 1275); χ2, Chi-square (non-significant values suggest good fit); RMSEA, root-mean-square error of approximation (cutoffs of .01, .05, and .08 indicate excellent, good, and acceptable fit, respectively3); TLI=Tucker-Lewis index4 (≥.95 indicates good fitting models5); CFI, Bentler’s comparative fit index6 (≥.96 indicates goodness of fit)

**eTable 7: Group means of symptom changes and number of services lost and modified in the aggregate dataset and by COVID19 impact subgroups.**

| Characteristic | **Aggregate** | **S1** | **S2** | **S3** | **S4** |  | **ANOVA Subgroup comparisons** | | |
| --- | --- | --- | --- | --- | --- | --- | --- | --- | --- |
|  | N=1275 | (n=251,20) | (n=653,51) | (n=293, 23%) | (n=78, 6%) | *F (df1-df2)* | *P-value adj.* | **η^2^** | Post Hoc comparisons |
| **Symptom Domain Change (Current-Prior), M (SD)^a^** |  |  |  |  |  |  |  |  |  |
| Adaptive Living Skills | 0.3 (1.6) | 1.7 (2) | -0.1 (1.3) | 0 (1.3) | -0.3 (1.2) | 102.9 (3-1271) | P<0.001 | 0.20 | S1 > S3 = S2 = S4 |
| RRB-LO | -0.1 (2.1) | 1.4 (2.5) | -0.5 (1.7) | -0.5 (1.8) | -0.5 (2.1) | 67.4 (3-1271) | P<0.001 | 0.14 | S1 > S3 = S2 = S4 |
| RRB-HO | -0.1 (1.6) | 0.8 (1.7) | -0.2 (1.6) | -0.2 (1.2) | -0.7 (1.5) | 33.5 (3-1271) | P<0.001 | 0.07 | S1 > S3 = S2 = S4 |
| Activity/Inattention | 0.1 (1.8) | 1.4 (2) | -0.3 (1.6) | -0.1 (1.4) | -0.3 (1.6) | 75.1 (3-1271) | P<0.001 | 0.15 | S1 > S3 = S2 = S4 |
| Oppositional | 0.3 (3) | 2.9 (3.7) | -0.5 (2.6) | 0.2 (1.7) | -0.8 (2.5) | 104.6 (3-1271) | P<0.001 | 0.20 | S1 > S3 > S2 = S4 |
| Anxiety | 0.3 (2.8) | 2.9 (3.6) | -0.6 (2.1) | 0.2 (1.8) | -0.8 (2.5) | 130.5 (3-1271) | P<0.001 | 0.24 | S1 > S3 > S2 = S4 |
| Sleep Problems | 0.2 (1.5) | 1.7 (1.8) | -0.2 (1.2) | -0.1 (1.1) | 0.2 (1.4) | 122.4 (3-1271) | P<0.001 | 0.22 | S1 > S3 = S2 = S4 |
| **Total Raw Number of Services, M (SD)** |  |  |  |  |  |  |  |  |  |
| Lost School Services | 1.2 (1.7) | 1.3 (1.7) | 1.0 (1.3) | 0.5 (0.8) | 5.3 (1.5) | 279.5 (3-1271) | P<0.001 | 0.40 | S4 > S1 = S2 > S3 |
| Lost Out. School Services | 1.3 (2) | 1.4 (1.9) | 1.0 (1.5) | 0.5 (1) | 6.1 (1.7) | 311.1 (3-1271) | P<0.001 | 0.42 | S4 > S1 = S2 > S3 |
| Continued School Services | 1.1 (1.4) | 0.7 (1.1) | 0.5 (0.8) | 3 (1.7) | 0.3 (0.6) | 306.9 (3-1271) | P<0.001 | 0.42 | S3 > S1 = S2 = S4 |
| Continued Out. School Services | 0.8 (1.3) | 0.6 (0.8) | 0.4 (0.7) | 2.1 (1.7) | 0.3 (0.5) | 193.8 (3-1271) | P<0.001 | 0.31 | S3 > S1 = S2 = S4 |
| ^a^Aggregate and subgroup scores are raw difference scores from each domain, shown as M (SD).Abbreviations: S1, Broad symptom worsening only subgroup; S2, Average symptom/service changes subgroup; S3, Primarily modified services subgroup; S4, Primarily lost services subgroup; RRB-LO, Restricted and Repetitive Behaviors - Lower Order; RRB-HO, Restricted and Repetitive Behaviors - Higher Order; Out., Outside; df1-df2,numerator and denominator degrees of freedom, P-value adj., P-values adjusted for FDR-correction (⍺=.05); η2, eta squared effect size | | | | | | | | | |

**eTable 8: Group means of top-ranked predictors identified via random forest analyses in the aggregate dataset and by subgroups.**

| **Random Forest top-ranked predictors, M (SD)** | **Aggregate** | **S1** | **S2** | **S3** | **S4** | **ANOVA Subgroup comparisons** | | | |
| --- | --- | --- | --- | --- | --- | --- | --- | --- | --- |
|  | N=1244 | (n=249,20%) | (n=637, 51%) | (n=283,23%) | (n=75,6%) | *F (df1-df2)* | *P-value adj.* | **η^2^** | Post Hoc comparisons |
| Pre-pandemic School Services, total services^a^ | 2.3 (2) | 1.9 (1.8) | 1.5 (1.5) | 3.4 (1.8) | 5.7 (1.4) | 196.208 (3, 1240) | P<0.001 | 0.322 | S4 > S3 > S1 > S2 |
| Pre-pandemic Outside of School Services, total services^a^ | 2 (2) | 1.8 (1.8) | 1.5 (1.6) | 2.4 (1.9) | 6.4 (1.6) | 195.063 (3, 1240) | P<0.001 | 0.321 | S4 > S3 > S1 > S2 |
| Stringency Index, raw score [range:0-100] | 62.8 (12) | 62.8 (12.3) | 61.5 (11.5) | 66.8 (12.5) | 59 (9.2) | 16.188 (3, 1240) | P<0.001 | 0.038 | S3> (S1 = S2 = S4) |
| Lifestyle stress, raw score [range:2-10] | 5.7 (2.3) | 6.7 (2.3) | 5.3 (2.2) | 5.5 (2.2) | 6.2 (2.4) | 25.95 (3, 1240) | P<0.001 | 0.059 | S1 = S4>(S3 = S2) |
| COVID-19 worries, raw score [range:4-20] | 8.5 (3.6) | 9.9 (4.2) | 8.1 (3.3) | 8.3 (3.2) | 7.9 (4) | 18.03 (3, 1240) | P<0.001 | 0.042 | S1 >(S3 = S2 = S4) |
| New COVID-19 infections, new cases/day [range:0-8460] | 446.7 (1009.9) | 364.1 (880.4) | 396.6 (989.2) | 723 (1215.5) | 104.4 (199.6) | 11.282 (3, 1240) | P<0.001 | 0.027 | S3 >(S2 = S1 = S4) |
| Child age, years [range:5-21] | 11 (3.6) | 10.8 (3.6) | 11.6 (3.5) | 10.2 (3.4) | 9.7 (3.1) | 15.842 (3, 1240) | P<0.001 | 0.037 | S2 >(S1 > S4 = S3) |
| Pre-pandemic Global Severity, z score^b^ | 0 (0.6) | 0 (0.5) | -0.1 (0.6) | -0.1 (0.6) | 0.3 (0.7) | 9.544 (3, 1240) | P<0.001 | 0.023 | S4>(S1 = S3 = S2) |
| Random Forest top-ranked predictors above are shown in descendent order of feature importance. ^a^Total number of services that the child was receiving prior to the COVID-19 pandemic. In school settings ranging from 0 to 7 total services; and in outside school settings, ranging from 0 to 8 total services. ^b^Pre-pandemic Global severity is an averaged z score, its calculation is described in Methods in the Supplement. Abbreviations: S1, Broad symptom worsening only subgroup; S2, Average symptom/service changes subgroup; S3, Primarily modified services subgroup; S4, Primarily lost services subgroup df1-df2, numerator and denominator degrees of freedom, P-value adj., P-values adjusted for FDR-correction (⍺=.05); η2, eta squared effect size | | | | | | | | | |

**eFigure 1. CRISIS AFAR Parent Baseline Survey v0.5.1 (3-21 years): Domain Structure and items.**

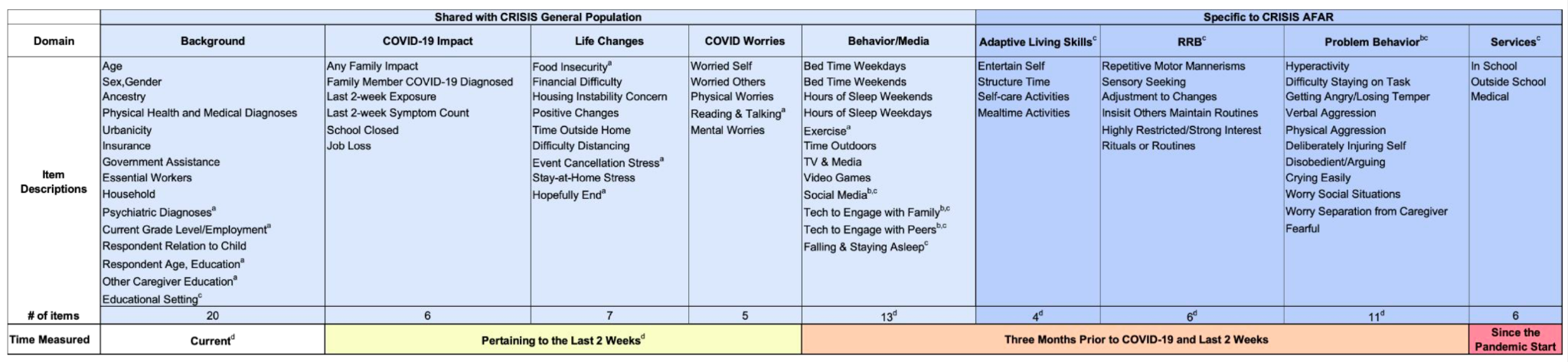

CRISIS AFAR retained CRISIS key content (light blue sections) and structure (last 2 weeks for COVID-19 impact, COVID-19 worries and life changes domains, the 3 months prior to the pandemic and last 2 weeks for the other Likert-scale for behavior questions) and newly included specific assessment domains for behavior and service changes (dark blue sections). **^a^** Item slightly reworded; ^b^ Item skipped for children younger than 5 years; ^c^ Item added in CRISIS AFAR. **^d^** Items asked twice for 3 months prior and to the pandemic and last 2 weeks. See Methods in eAppendix for more details on the adaptation process. RRB: Restricted Repetitive Behaviors/Interests.

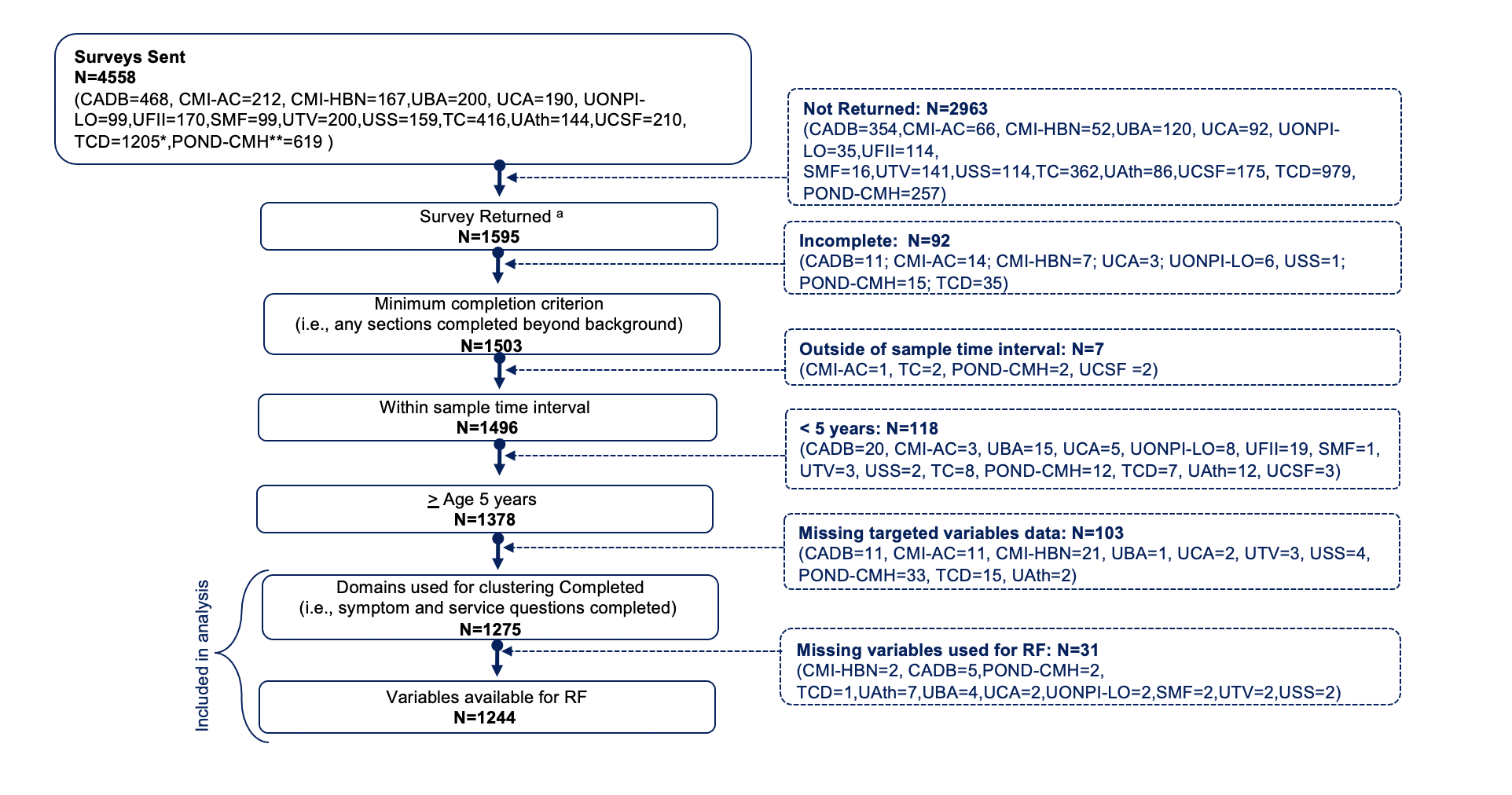
**eFigure 2. Data collection and selection flow.**

Across 14 institutions (15 samples) 4558 families are known to be contacted to complete the survey. Of them, a total of N=1595 (36%) CRISIS AFAR baseline parent-based surveys were returned with return rate varying by sample (CADB=114, 24%; CMI-AC=146, 69%; CMI-HBN=115, 69%: UBA=80, 40%; UCA=98, 52%; UONPI-LO=64, 65%;UFII=56 ,33%; SMF=83, 84%,UTV=59 ,30%;USS=45,28%;TC=53,13%; UAth=58,40%; UCSF=35,17%; TCD*=226,19%; POND-CMH**=362,58% . Of those returned, N=1503 of which were completed for at least two sections including background. For data analyses we selected those completed survey within 2 weeks from the completion time interval of 90% of a given sample (N=1496), of children aged 5 years-old and above (N=1378) who responded to AFAR behavioral and service domains examined with hierarchical clustering. This yielded a final aggregate dataset of N=1275 across the 15 contributing samples. See Figure 1 for site name abbreviated labels. *For TCD sample, the total number of surveys sent for one of the sources is unknown. **For POND-CMH sample the original contact count includes a larger group of children than those with NDD targeted here. Abbreviations: see eTable 1 for site name abbreviated labels.

**eFigure 3. CRISIS AFAR Survey background and COVID-related information for the aggregate and each contributing sample.**

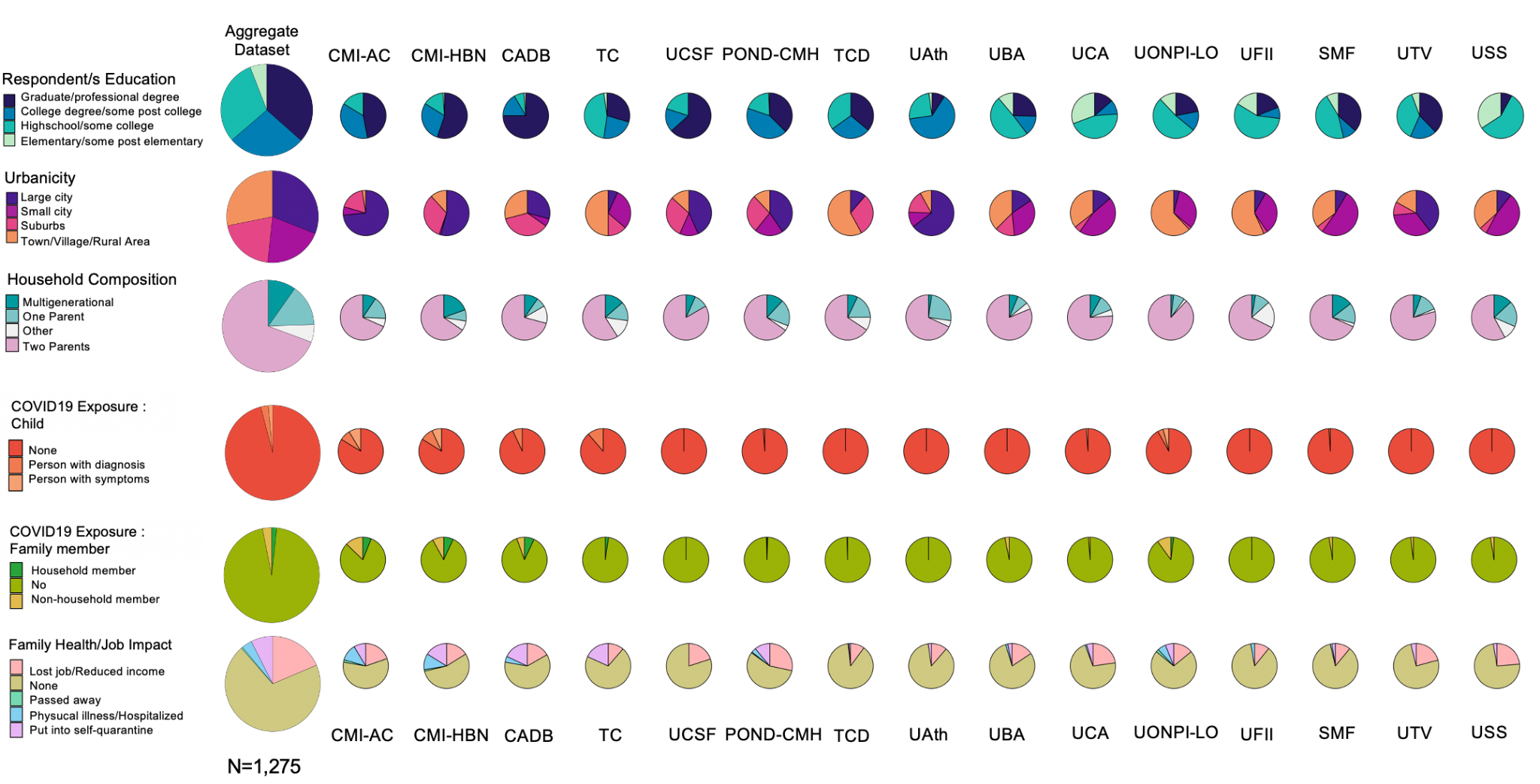

Larger pies show the aggregate dataset information (N=1275), and smaller pies each of the contributing samples. Of them, the first 4 samples from the right include those collected in USA institutions (CMI-AC, CMI-HBN, CADB, TC and UCSF), followed by the one collected in Canada (POND-CMH), Ireland (TCD), and Greece (UAth), and the f7 samples from Italian institutions (UBA, UCA, UONPI-LO, UFII, SMF, UTV, USS). See Figure 1 for each sample label. The first 3 rows refer to the background survey questions: highest level of respondent education (first or second caregiver), urbanicity and household composition; the 3 bottom rows illustrate COVID-19 related factors: child and family member exposure as well as family impact on health and job. Abbreviations: see eTable 1 for site name abbreviated labels.

**eFigure 4. Symptom and service access change pattern**

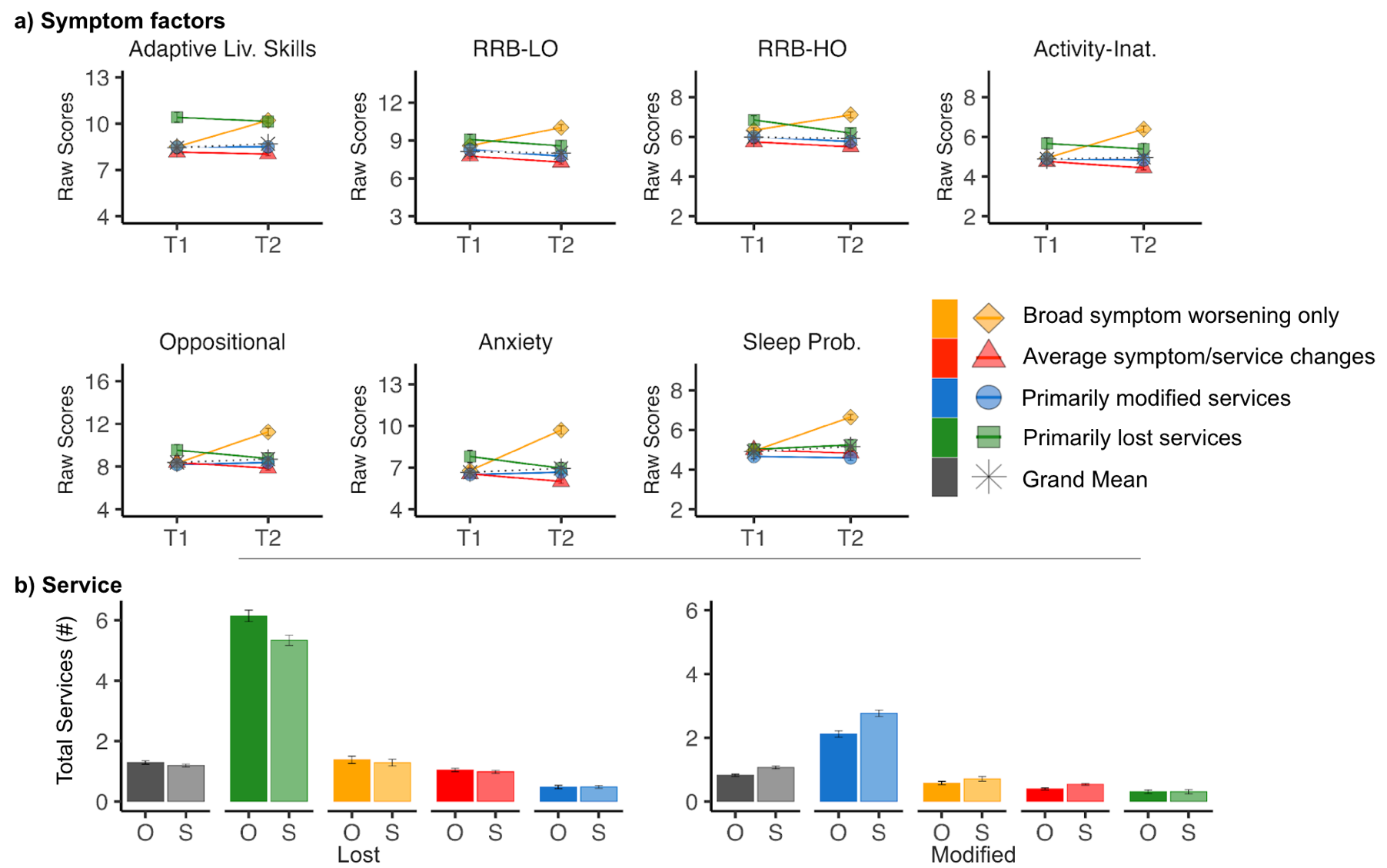

a) Mean and standard errors bars of prior (T1) and current (T2) symptom scores for each of the seven factors examined in clustering analyses for each subgroup (color coded) and for the aggregate sample (in gray).  For each graph the y axis is scaled using the minimum and maximum scores of each symptom factor. Of note, the scores from adaptive living skills domain were rescaled to follow the same scale as the other factors. For each factor higher values in the y axis indicate higher severity of symptoms. b) Mean number of services lost or modified (and their standard error bars) for each subgroup (color coded), as well as the aggregate dataset (gray bars) at school (S; light colored bars) or outside school (O; darker bars).

**eFigure 5. Subgroup distribution by contributing sample.**

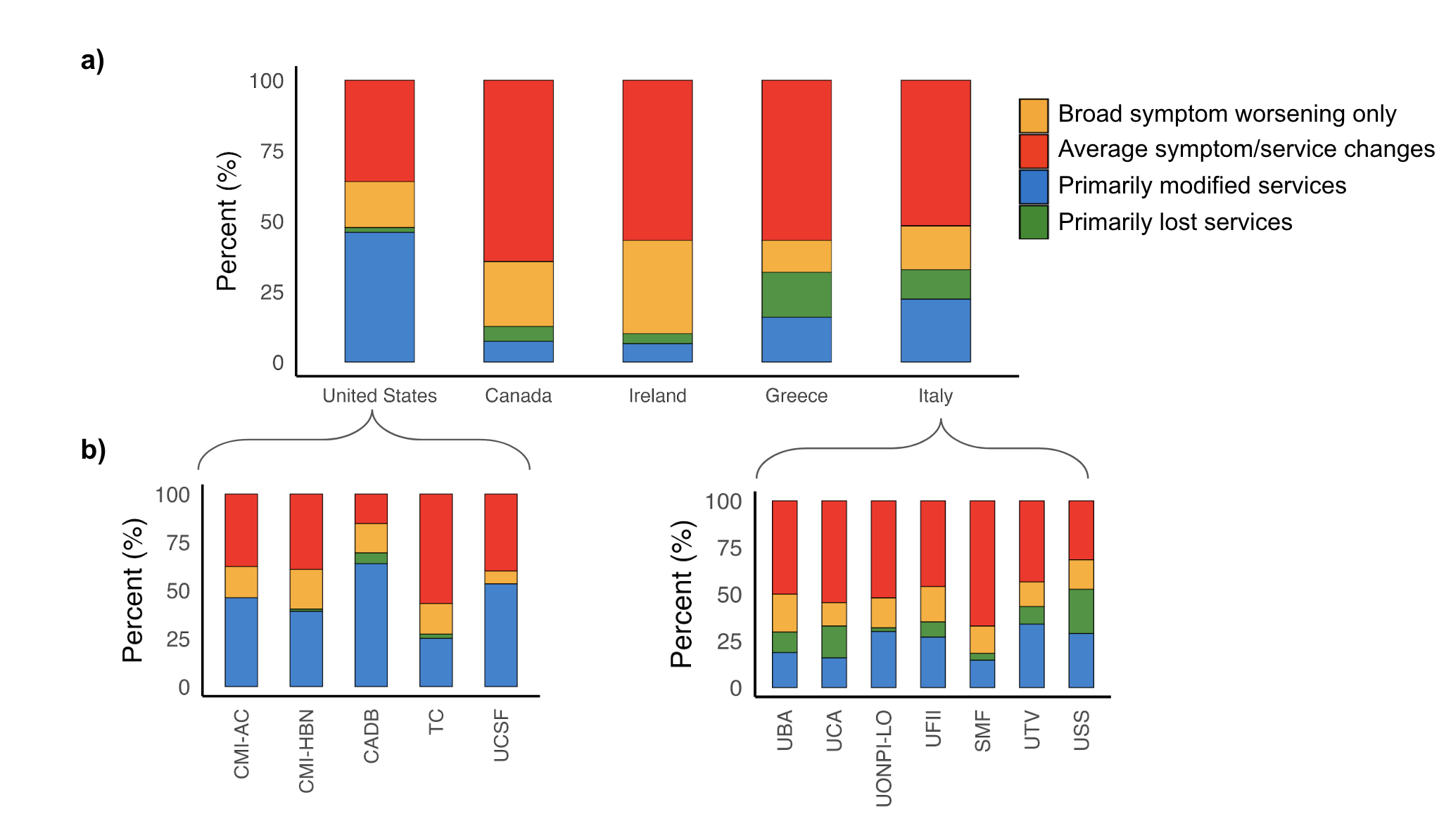

The percentages of children for each subgroup based on the total aggregate dataset (N=1275) by Nation are shown (n=5: United States, Canada, Ireland, Greece, and Italy (a) and contributing sample in the United States (left, n=5) and Italy (right, n=7) (b). Abbreviations: see eTable 1 for site name abbreviated labels.

**eFigure 6. Correlations between pre-pandemic standardized severity measures and AFAR baseline global severity score.**

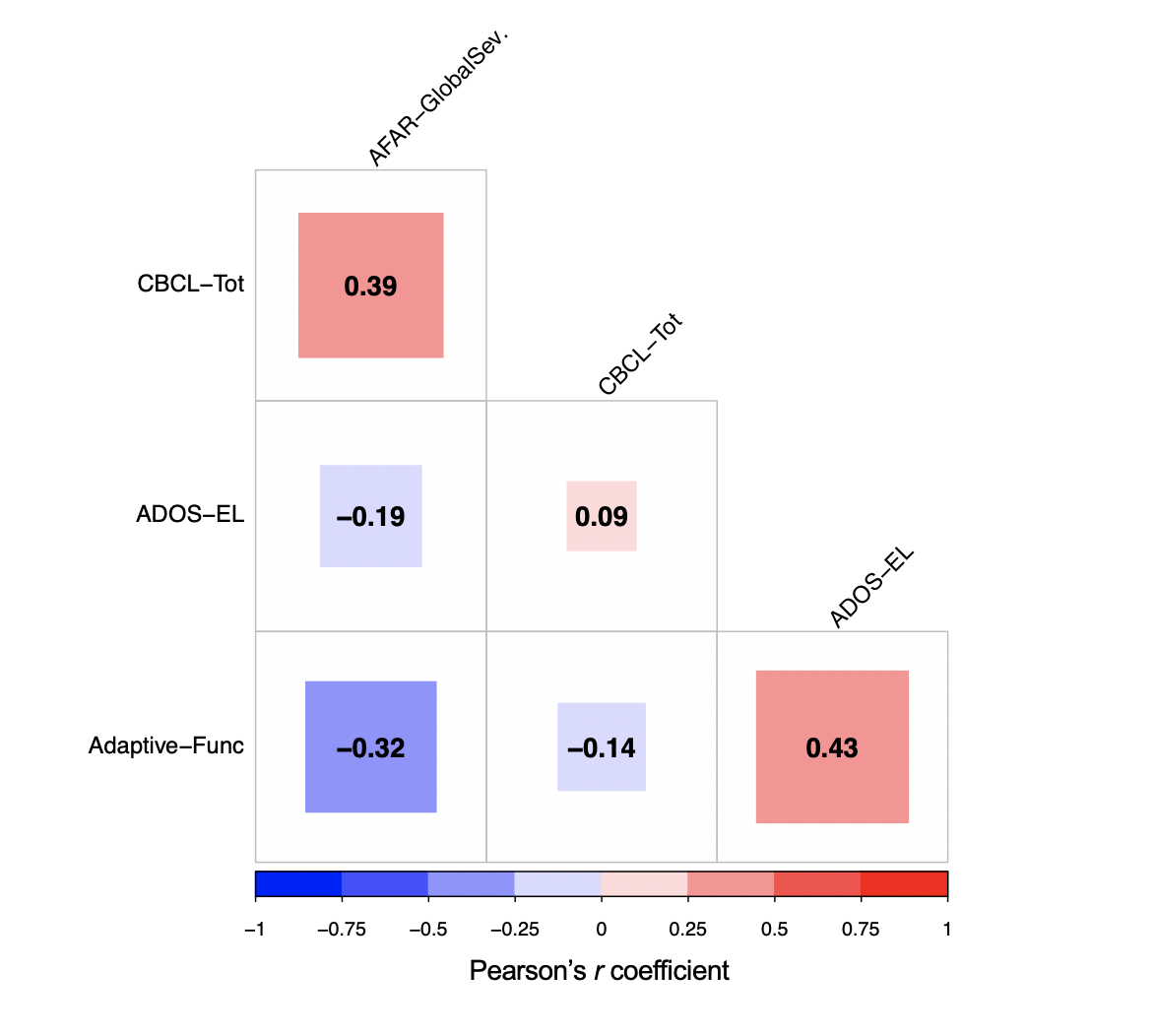

Correlations were calculated across N=453 children with complete observations from 9 contributing samples. The size of the squares within the matrix is proportional to the Pearson’s r coefficient values (i.e., larger squares indicate stronger correlations). The color scale indicates whether the correlation is positive (in red) or negative (in blue). Of note, all correlations with AFAR global severity scores (“AFAR-GlobalSev.”) are statistically significant at p<0.0001 after FDR-correction. Abbreviations: AFAR-GlobalSev., global severity prior to the COVID-19 pandemic (see Methods in eAppendix for details on its calculation); CBCL-Tot, CBCL T total scores; ADOS-EL, ADOS-2 Expressive language scores; Adaptive-Func, composite scores of adaptive functioning.

10.	American Psychiatric Association. *Diagnostic and Statistical Manual of Mental Disorders*. American Psychiatric Association; 2013.

11.	Guttentag S, Bishop S, Doggett R, et al. The Utility of Parent-Report Screening Tools in Differentiating Autism vs. ADHD in School-age Children. doi:10.31234/osf.io/9pu7t

24.	Association AP, Others. DSM-4-TR. Published online 2004.

25.	American Psychiatric Association DS, Association AP, Others. *Diagnostic and Statistical Manual of Mental Disorders: DSM-5*. Vol 5. American psychiatric association Washington, DC; 2013.

26.	World Health Organization. *The International Statistical Classification of Diseases and Health Related Problems ICD-10: Tenth Revision. Volume 2: Instruction Manual*. World Health Organization; 2004.

27.	Rutter M, Le Couteur A, Lord C, Others. Autism diagnostic interview-revised. *Los Angeles, CA: Western Psychological Services*. 2003;29(2003):30.

28.	Lord C, Risi S, Lambrecht L, et al. The Autism Diagnostic Observation Schedule—Generic: A Standard Measure of Social and Communication Deficits Associated with the Spectrum of Autism. *J Autism Dev Disord*. 2000;30(3):205-223.

29.	Lord C, Rutter M, DiLavore P, et al. Autism diagnostic observation schedule--2nd edition (ADOS-2). *Los Angeles, CA: Western Psychological Corporation*. 2012;284.

30.	Kaufman J, Birmaher B, Brent D, et al. Schedule for Affective Disorders and Schizophrenia for School-Age Children-Present and Lifetime Version (K-SADS-PL): initial reliability and validity data. *J Am Acad Child Adolesc Psychiatry*. 1997;36(7):980-988.

31.	Sparrow SS, Cicchetti D, Saulnier C. Vineland adaptive behavior scales--third edition (Vineland-3). *Circle Pines, MN: American Guidance Service*. Published online 2016.

32.	Harrison PL, Oakland T. *ABAS-3*. Western Psychological Services Torrance; 2015.

39.	Maechler M, Rousseeuw P, Struyf A, Hubert M, Hornik K. Cluster: Cluster Analysis Basics and Extensions.(2021). R package version 2.1. 2—For new features, see the ‘Changelog’file (in the package source).
